# Iron deficiency in Women of Childbearing Age with Self-reported Oral Iron Gastrointestinal Intolerance and Management with an oral Iron-Whey-Protein Formulation

**DOI:** 10.1101/2021.09.01.21262983

**Authors:** JGF Gilmer, Fiona Ryan, Anna Seoighe, Maria Jose Santos-Martinez, Cristin Ryan, Mark Ledwidge

## Abstract

**Background:** Intolerance to oral iron is thought to result in poor adherence and persistence of nutritional deficit amongst women of childbearing age, however few studies have evaluated oral iron intolerance, iron deficiency and anaemia in this setting. Iron-whey protein microspheres (IWP) could help.

**Methods:** We documented self-reported oral iron gastrointestinal intolerance, ferritin and haemoglobin levels in a screening study of women of childbearing age. Following a washout period of 16 days, we randomised 59 of these women with iron deficiency, stratified according to the presence of anaemia, to three doses of IWP: (14mg daily, 25mg daily and 50mg daily). We excluded those with established gastrointestinal disease, potential allergy to whey protein and severe anaemia. The primary endpoint was persistence and adherence (>80% based on pill-counts). Secondary endpoints included changes in self-reported oral iron gastrointestinal intolerance, gastro-intestinal symptom rating scale (GSRS), serum iron, serum ferritin, transferrin saturation and haemoglobin levels.

**Results:** A total of 128 (62.7%) of the participants had low iron stores (ferritin < 30 µg/L), 65 (31.9%) had moderate to severe iron deficiency (ferritin <12 µg/L) and 33 (16.2%) had iron deficiency anaemia. Amongst 59 women who participated in the prospective study, 48 (81.4%) were classified as adherent/persistent with therapy using IWP compared to 12 (20.3%) taking the prior oral iron p<0.0001. These patients also showed significantly fewer reports of gastrointestinal intolerance with IWP (0.59 ± 0.91) and lower GSRS scores (6.2 ± 7.5) compared to the previous oral iron product (3.98 ± 2.22, and 15.6 ± 9.7 respectively, both P<0.0001). There were no differences in adherence, self-reported adverse GI effects and GSRS between the dose groups during the study. Serum iron levels increased across the whole cohort from 11.3 ± 7.4 μmol/L to 20.5± 11.0 μmol/L (P<0.0001), transferrin saturation levels increased from 18.4 ± 13.3 % to 33.6 ± 17.6 % (P<0.0001) and median ferritin levels overall increased from 8.00 [IQR 6.00;13.0] to 15.5 [IQR 9.00;24.2] µg/L at 12 weeks (P=0.0002). Haemoglobin levels increased from 11.36 g/dL (95%CI 10.95 to 11.77) to 12.40 g/dL (95%CI 12.03 to 12.76, P=0.0007) in patients with anaemia and were normalised in most patients taking 50mg IWP daily.

**Conclusions:** Low iron, iron deficiency and anaemia are common in women of childbearing age with a history of intolerance to oral iron. Patients with low iron (ferritin < 30 µg/L) and moderate to severe iron deficiency (ferritin <12 µg/L) have similar impairment of energy. IWP can improve self-reported oral iron adherence and tolerability as well as iron stores, haemoglobin and tiredness in these women.

## BACKGROUND

Pre-menopausal adult women are at high-risk of low iron stores, iron deficiency and anaemia because of inadequate iron intake and menstrual blood loss.[1,2] This is frequently managed using oral iron supplementation. However, due to low fractional absorption of oral iron, [3] high doses (e.g. ferrous sulfate at 100-200mg elemental iron daily) are commonly used, causing adverse gastrointestinal (GI) effects in a majority of patients.[4] This results in poor adherence in up to 50% of patients and continuation of nutritional deficit.[5]

Despite widespread recognition of this problem, there are few data on the prevalence of low iron stores (ferritin < 30 µg/L) in consecutive, consenting, adult women of childbearing age with self-reported adverse oral iron gastrointestinal (GI) effects. Furthermore, available data in this setting have advocated intravenous iron infusions, which require resource-intensive administration as well as monitoring in a healthcare setting.[6] Adverse GI effects with oral iron are attributed to damage to the intestinal mucosa, in part due to oxidative stress.[7-9] Adverse effects are dose related [10,11]. Symptoms can be characterised as upper GI (e.g. nausea, abdominal pain, bloating, eructation) or lower GI (constipation, diarrhoea), although sufferers frequently report both [11]. While enteric coated or delayed release formulations of oral iron can, in principle, address upper GI adverse effects, they can also reduce absorption and potentially aggravate lower GI effects, further compromising adherence and absorption.[12,13] A systematic review of oral iron treatment studies concluded that there is much heterogeneity in the reporting of adverse GI symptoms associated with use of oral iron. It found specific evidence for increased constipation, abdominal pain and diarrhoea with oral iron.[11] More studies are needed to understand the contribution of adverse GI effects to continuation of iron deficiency and related anaemia in adult women treated with oral iron. Furthermore, the validated Gastrointestinal Symptom Rating Scale (GSRS), which was developed for use in gastroenterology as a reliable screen for gastrointestinal disorders, could standardise reporting of a full range of adverse GI symptoms amongst oral iron users.[14-16]

We previously reported a formulation of ferrous iron in a de-calcified, denatured whey protein (WP) matrix formulation at a daily elemental iron dose of 25mg daily.[17] The formulation improves iron absorption and also results in less iron induced oxidative stress in Caco-2 and HT29 intestinal epithelial cell lines *in-vitro* than other presentations of iron tested under similar conditions. This suggests the formulation may have value in managing iron deficiency associated with oral iron intolerance, although to date there are no prospective, comparative clinical data on different doses over time.

The present study aims to firstly investigate the prevalence of low iron stores, iron deficiency and anaemia in a screening cohort study of consecutive, consenting, adult women of childbearing age with a self-reported history of intolerance to oral iron. Secondly, it prospectively compares the 12-week adherence, GI tolerability and efficacy associated with three different doses of an Iron-Whey Protein formulation (IWP) in this difficult to treat cohort.

## METHODS

### Participants

The first part of this investigation involved a screening cohort study evaluating the prevalence of low iron stores (ferritin <30µg/L), iron deficiency (ferritin < 12µg/L) and anaemia (haemoglobin <12 g/dL) amongst adult women of childbearing age with self-declared oral iron GI intolerance. Consecutive, pre-menopausal, adult women (18-55 years) with a self-reported history of gastrointestinal intolerance to oral iron and no other diagnosed gastrointestinal disease (current inflammatory bowel disease, including Crohn’s disease and ulcerative colitis, as well as irritable bowel disease) were invited to a clinic visit where they were evaluated for self-reported adverse GI effects, iron biochemistry and haemoglobin. For consenting patients with ferritin <30 µg/L, who were otherwise considered generally of good health, were willing to comply with the screening protocol and were willing to undergo a two-week washout of their previous oral iron, the preliminary study was followed by a randomised, prospective, double-blind, parallel group, dose-finding, clinical study. This study tracked changes in iron stores, haemoglobin, self-reported adverse GI effects, GSRS and adherence from the previous oral iron to IWP at three different elemental iron doses over 12 weeks. Excluded were patients taking concurrent medication which interferes with the absorption of iron (e.g. tetracyclines, calcium supplements), those with a history of dairy allergy or were hypersensitive to any of the components of the test product, those with severe anaemia (females with haemoglobin <9.5 g/dL and a malignant disease or any concomitant end-stage organ disease or significant acute or chronic, unstable and untreated disease or any condition, which contraindicated, in the investigator’s judgement, entry to the study.

### Intervention and main outcome measures

Included patients with ferritin < 30 ug/L were randomised to IWP (Active Iron®) in one of three groups: a standard 25mg single daily dose in the morning with matching dummy capsule in the evening, a lower (14mg) single daily dose in the morning and matching dummy capsule in the evening or twice daily with 25mg (50mg daily). Patients were analysed in pre-specified stratified subgroups with and without anaemia. The primary endpoint of the prospective randomised controlled study was the change in proportion of subjects adherent and persistent (>80% based on pill counts) averaged from baseline to weeks 6 and baseline to week 12. Secondary endpoints were the change in self-reported upper and lower GI tolerability, GSRS, ferritin, transferrin saturation and haemoglobin over 12 weeks. The study was approved by the Cork University Hospital Research Ethics Committee and conformed to the principles of the Declaration of Helsinki and all patients provided written, informed consent. Detailed study procedures are presented in the Supplemental File.

### Statistical Analyses

All analyses was carried out using R version 4.0.1 (2020). Descriptive data are presented as n (%) as well as either mean ± SD or median (25th:75th percentile) for normally and non-normally distributed continuous variables, respectively. Shapiro-Wilk’s test used to formally assess normality of the variable data. Frequencies and percentages (in parentheses) summarize categorical variables. If continuous variables were transformable to normal, they were power-transformed and independent, two sample t-tests were used for analysis of continuous variables. If data were not transformable to normal, non-parametric t-test equivalents (Wilcoxon signed rank and rank sum test, Mann-Whitney test and analysis of covariance [ANCOVA]) were used. Chi-squared (or Fisher Exact) analyses were used to compare categorical variables as appropriate. Repeated marker changes from baseline to 12 weeks (ferritin, haemoglobin, transferrin saturation) were also analysed using ANOVA with repeated measures models. Primary and secondary outcome measures were performed both with and without adjustment for the effects of baseline age, body mass index and systolic blood pressure. Further models included adjustment for pre-specified baseline outcomes of interest. Categorical endpoints were analysed using generalized linear modelling with a binomial outcome distribution for prevalence. A p-value of ⍰ 0.05 was considered statistically significant.

## Results

### Screening Cohort Study

In the screening cohort study, the main finding was that of 204 consecutive participating adult women of childbearing age, with a history of gastrointestinal intolerance to oral iron, almost two in three had low iron stores or iron deficiency (ferritin <30 µg/L, n=128, 62.7%). A total of 33 (16.2%) also had anaemia (haemoglobin <12 g/dL). Surprisingly, only 17 (8.3%) had a self-reported history of iron deficiency and 26 (16.9%) reported prior iron deficiency or anaemia. Furthermore, there was no univariate or multivariable association between a self-reported history of iron deficiency and low iron stores (multivariable odds ratio 1.10, 95% CI 0.85 - 1.45). Overall, the prevalence of iron deficiency was much higher than expected and the majority of subjects did not recall a formal diagnosis of iron deficiency or anaemia.

Moderate to severe iron deficiency (ferritin cut-off 12µg) affected 65 (31.9%) women; 24 (11.8%) of these also had anaemia. Unlike the low iron stores cut-off, there was a significant univariate and multivariable association between prevalent iron deficiency using the ferritin cut-off of 12µg/L and a self-reported history of iron deficiency (multivariable odds ratio 1.41, 95% CI 1.12- 1.78, p=0.004) as well as a history of anaemia (multivariable odds ratio 1.31, 95% CI 1.07- 1.60, p=0.011). Complete blood counts were available from the study in 123 (60.3%) of the total cohort and similar levels of low iron stores was evident in this subgroup (n=82, 66.7%). In this subgroup, low iron stores were strongly and independently associated with lower parameters of functional and storage iron (serum iron, iron binding capacity, transferrin saturation), lower red blood cell indices (haemoglobin, haematocrit, mean corpuscular haemoglobin, mean corpuscular haemoglobin concentration) and higher red blood cell distribution width (Supplemental File Table S1).

Overall, the participants in the screening study reported their experience of adverse GI effects with oral iron products in the following way: lower-GI tract intolerance occurred more often (n=186, 91.2%) than upper-GI tract intolerance (n=136, 66.7%, p <0.01, Table 1). Most women (n=119, 58.3%) reported combined upper and lower GI tract symptoms. The single most common oral iron adverse GI effect reported by the cohort was constipation (affecting almost 8 out of 10 women). This was followed by abdominal pain (approx. 4 in 10), then nausea (3 in 10, Table 1). Combined abdominal pain and nausea was reported by 118 participants (57.8%). There was no apparent association between the profile of oral iron intolerance and ferritin < 30µg/L. Although we observed univariate differences in the profile of GI intolerance between those with ferritin levels above and below a cut-off of 12µg/L, none of these was significant when adjusted for age, body mass index, systolic blood pressure. Interestingly, we observed a significantly higher rate of self-reported upper GI intolerance in patients with moderate to severe iron deficiency using a threshold of 12µg/L (Supplemental File Table S1), which remained significant in multivariable analysis (adjusted odds ratio, 1.20, 95% CI 1.05 - 1.38, p<0.01).

**Table 1.** Demographic, anthropomorphic, haematinic and self-reported oral iron gastrointestinal tolerability profile of adult women of childbearing age with ferritin <30µg/L and a self-reported gastrointestinal intolerance to oral iron. Abbreviations: GI = gastrointestinal; SBP = systolic blood pressure; DBP = diastolic blood pressure; HR = heart rate; bpm = beats per minute; BMI = body mass index; Hb = haemoglobin.

|  | All Participants<br>N=204 | Normal Ferritin<br>N=76 | Ferritin < 30 µg/L<br>N=128 | P<br>value |
| --- | --- | --- | --- | --- |
| Age, years | 36.6 (10.1) | 36.8 (10.0) | 36.6 (10.2) | 0.911 |
| History of iron deficiency, n(%) | 17 (8.33%) | 4 (5.26%) | 13 (10.2%) | 0.337 |
| History of anaemia, n(%) | 23 (11.3%) | 6 (7.89%) | 17 (13.3%) | 0.249 |
| History of iron deficiency or anaemia, n(%) | 26 (12.7%) | 8 (10.5%) | 18 (14.1%) | 0.606 |
| Weight, kg | 70.7 [60.7;81.4] | 70.4 [60.9;83.1] | 71.3 [60.6;79.7] | 0.784 |
| Height, m | 1.65 (0.06) | 1.65 (0.06) | 1.65 (0.06) | 0.756 |
| BMI, kg/m<sup>2</sup> | 25.8 [22.3;29.6] | 25.5 [22.3;30.4] | 25.9 [22.2;29.3] | 0.962 |
| Temp, °C | 36.3 (0.47) | 36.3 (0.47) | 36.2 (0.46) | 0.064 |
| SBP, mmHg | 109 [102;117] | 109 [102;117] | 109 [102;117] | 0.900 |
| DBP, mmHg | 73.8 (9.38) | 73.8 (8.24) | 73.9 (10.0) | 0.923 |
| HR, bpm | 70.0 [65.0;77.0] | 69.5 [64.0;76.0] | 70.0 [65.0;77.0] | 0.640 |
| Smoking status: |  |  |  | 0.890 |
| Never smoked, n(%) | 133 (65.2%) | 49 (64.5%) | 84 (65.6%) |  |
| Previous smoker, n(%) | 47 (23.0%) | 17 (22.4%) | 30 (23.4%) |  |
| Current smoker, n(%) | 24 (11.8%) | 10 (13.2%) | 14 (10.9%) |  |
| Alcohol consumption, units/week | 2.64 (2.96) | 2.91 (3.25) | 2.48 (2.77) | 0.345 |
| Depot contraceptive, n(%) | 8 (3.92%) | 4 (5.26%) | 4 (3.12%) | 0.474 |
| Patch or ring contraceptive, n(%) | 15 (7.35%) | 9 (11.8%) | 6 (4.69%) | 0.106 |
| Oral contraceptive, n(%) | 37 (18.1%) | 15 (19.7%) | 22 (17.2%) | 0.788 |
| Serum Iron, µmol/L | 14.7 [9.25;21.4] | 18.8 [15.8;26.9] | 12.7 [7.18;17.1] | <0.001 |
| Total iron binding concentration, µmol/L | 61.0 (9.94) | 54.4 (8.73) | 64.0 (8.96) | <0.001 |
| Transferrin saturation, % | 25.7 [13.7;35.5] | 36.2 [28.8;50.7] | 21.2 [9.85;29.0] | <0.001 |
| Ferritin, µg/L | 18.0 [9.00;43.2] | 50.5 [41.0;66.0] | 11.5 [7.00;15.8] | <0.001 |
| White cell count, 10<sup>9</sup>/L | 5.60 [4.76;6.79] | 5.73 [5.17;7.05] | 5.58 [4.55;6.39] | 0.111 |
| Red cell count, 10<sup>12</sup>/L | 4.42 [4.24;4.70] | 4.43 [4.27;4.80] | 4.41 [4.18;4.66] | 0.418 |
| Hb, g/dL | 13.0 [12.2;13.7] | 13.5 [12.7;14.0] | 12.8 [11.9;13.6] | <0.001 |
| Haematocrit, L/L | 0.40 [0.37;0.42] | 0.41 [0.40;0.42] | 0.38 [0.36;0.41] | <0.001 |
| Mean cell volume, fL | 88.8 [85.4;92.1] | 91.9 [87.8;94.8] | 88.3 [84.3;90.4] | <0.001 |
| Mean cell Hb, pg | 28.9 [27.3;30.4] | 29.8 [28.5;31.3] | 28.5 [26.6;29.9] | <0.001 |
| Mean cell Hb concentration, g/dL | 32.4 (1.19) | 32.8 (1.20) | 32.2 (1.14) | 0.009 |
| Red cell distribution width, % | 13.3 [12.8;14.2] | 12.9 [12.6;13.4] | 13.6 [13.0;14.7] | <0.001 |
| Platelets, 10<sup>9</sup>/L | 287 [247;324] | 298 [270;324] | 278 [243;323] | 0.196 |
| Neutrophils, 10<sup>9</sup>/L | 3.26 [2.49;4.21] | 3.48 [2.83;4.47] | 3.20 [2.44;4.11] | 0.202 |
| Lymphocytes, 10<sup>9</sup>/L | 1.73 [1.36;2.06] | 1.84 [1.43;2.11] | 1.67 [1.33;1.88] | 0.046 |
| Monocytes, 10<sup>9</sup>/L | 0.44 [0.35;0.53] | 0.45 [0.34;0.56] | 0.44 [0.36;0.52] | 0.718 |
| Eosinophils, 10<sup>9</sup>/L | 0.14 [0.08;0.26] | 0.16 [0.10;0.27] | 0.12 [0.06;0.25] | 0.076 |
| Basophils, 10<sup>9</sup>/L | 0.03 [0.02;0.04] | 0.03 [0.02;0.05] | 0.02 [0.02;0.04] | 0.218 |
| History of GI disease, n(%) | 17 (8.33%) | 5 (6.58%) | 12 (9.38%) | 0.662 |
| Constipation, n(%) | 163 (79.9%) | 63 (82.9%) | 100 (78.1%) | 0.521 |
| Abdominal pain, n(%) | 81 (39.7%) | 27 (35.5%) | 54 (42.2%) | 0.428 |
| Nausea, n(%) | 60 (29.4%) | 19 (25.0%) | 41 (32.0%) | 0.365 |
| Abdominal pain or nausea, n(%) | 118 (57.8%) | 40 (52.6%) | 78 (60.9%) | 0.310 |
| Indigestion, n(%) | 27 (13.2%) | 9 (11.8%) | 18 (14.1%) | 0.811 |
| Heartburn, n(%) | 24 (11.8%) | 10 (13.2%) | 14 (10.9%) | 0.802 |
| Eructation, n(%) | 12 (5.88%) | 6 (7.89%) | 6 (4.69%) | 0.369 |
| Diarrhoea, n(%) | 12 (5.88%) | 4 (5.26%) | 8 (6.25%) | 1.000 |
| Vomitting, n(%) | 4 (1.96%) | 0 (0.00%) | 4 (3.12%) | 0.299 |
| Any lower adverse GI effects, n(%) | 186 (91.2%) | 71 (93.4%) | 115 (89.8%) | 0.538 |
| Number lower adverse GI effects | 1.00 [1.00;2.00] | 1.00 [1.00;1.00] | 1.00 [1.00;2.00] | 0.813 |
| Any upper adverse GI effects, n(%) | 136 (66.7%) | 45 (59.2%) | 91 (71.1%) | 0.112 |
| Number upper adverse GI effects | 1.00 [0.00;2.00] | 1.00 [0.00;1.25] | 1.00 [0.00;2.00] | 0.282 |
| Upper and lower adverse GI effects, n(%) | 119 (58.3%) | 40 (52.6%) | 79 (61.7%) | 0.260 |
| Only lower adverse GI effects, n(%) | 68 (33.3%) | 31 (40.8%) | 37 (28.9%) | 0.112 |
| Only upper adverse GI effects, n(%) | 17 (8.33%) | 5 (6.58%) | 12 (9.38%) | 0.662 |
| Number total adverse GI effects | 2.00 [1.00;3.00] | 2.00 [1.00;3.00] | 2.00 [1.00;3.00] | 0.244 |

### Prospective Randomised Controlled Treatment Study

Amongst the 128 women with oral iron gastrointestinal intolerance and ferritin < 30µg/L, 59 were eligible and agreed to participate in the double blind, prospective, randomised, controlled study of iron whey-protein microspheres (IWP). A total of 52 women were not included in the prospective treatment study because they had iron deficiency without anaemia and the stratified quota of 30 patients had already been reached. A total of 9 did not want to participate for personal reasons and 8 were excluded because of severe anaemia. Stratified inclusion resulted in the prospective treatment study participants (n=59) having lower ferritin and haemoglobin levels than the overall population of n=128 (Supplemental File Table S2), reflecting the stratified inclusion of similar numbers of women with and without anaemia. This subset was also more likely to have a history of iron deficiency and anaemia.

The baseline demographic, characteristics of this population in total and according to randomisation are presented in Table 2. The self-reported adverse GI effects and GSRS score related to previous oral iron products are presented in Table 3. The majority of the cohort (n=41, 69.5%) had previously been taking high dose (>60mg elemental Iron). A higher proportion of those previously taking > 60 mg elemental iron daily (n=34, 82.9%) reported constipation versus lower doses (<= 60mg daily, n=10, 55.6%, P=0.049), but otherwise there were similar numbers of adverse GI effects (4.07 ± 2.27 versus 3.78 ± 2.16 respectively, P=NS, Supplemental File Table S2). Women taking > 60 mg elemental iron of previous oral iron products also had lower haemoglobin levels, lower mean cell haemoglobin and higher red cell distribution width than those taking lower doses. As expected, there was a positive correlation between the number of self-reported oral iron adverse GI effects and the total GSRS score associated with taking those products, independent of age, blood pressure, heart rate and BMI. For each additional self-reported adverse GI effect, there was a 4.15-unit increase (95% CI 1.30-13.24, p=0.017) in GSRS associated with the previous oral iron product.

**Table 2.** Demographic, iron and full blood count profile of participants with ferritin <30µg/L and self-reported gastrointestinal intolerance to oral iron who were randomised to three different daily elemental iron doses of IWP (14mg, 25mg, 50mg). Abbreviations: IWP= iron-whey-protein formulation; SBP = systolic blood pressure; DBP = diastolic blood pressure; HR = heart rate; bpm = beats per minute; BMI = body mass index; Hb = haemoglobin.

|  | **All Patients**<br>**N=59** | **IWP 14mg**<br>**N=18** | **IWP 25mg**<br>**N=21** | **IWP 50mg**<br>**N=20** |
| --- | --- | --- | --- | --- |
| Age, years | 35.2 (11.0) | 35.3 (11.8) | 34.0 (10.0) | 36.1 (11.7) |
| SBP, mmHg | 109 [104;119] | 116 [108;123] | 105 [103;114] | 110 [100;122] |
| DBP, mmHg | 74.7 (9.49) | 75.3 (8.90) | 75.1 (8.34) | 73.7 (11.4) |
| HR, bpm | 70.3 (9.88) | 70.2 (10.4) | 70.6 (10.3) | 70.0 (9.47) |
| Weight, kg | 72.4 [59.6;82.4] | 75.5 [60.8;85.6] | 72.4 [62.4;78.2] | 69.7 [57.0;79.2] |
| BMI, kg/m<sup>2</sup> | 26.4 [22.0;30.6] | 27.1 [22.2;31.6] | 27.6 [22.6;30.2] | 25.1 [20.5;27.3] |
| Alcohol consumption, units/week | 3.55 (3.09) | 3.07 (2.94) | 4.40 (3.62) | 3.13 (2.72) |
| Smoking status: |  |  |  |  |
| Never smoked, n(%) | 9 (15.3%) | 1 (5.56%) | 5 (23.8%) | 3 (15.0%) |
| Previous smoker, n(%) | 40 (67.8%) | 15 (83.3%) | 13 (61.9%) | 12 (60.0%) |
| Current smoker, n(%) | 10 (16.9%) | 2 (11.1%) | 3 (14.3%) | 5 (25.0%) |
| Days since screening visit | 9.83 (3.90) | 10.1 (4.02) | 9.29 (2.90) | 10.2 (4.74) |
| Serum Iron, µmol/L | 9.60 [5.75;16.3] | 12.6 [5.73;18.5] | 10.3 [7.60;14.8] | 8.30 [5.45;14.5] |
| Unbound iron binding concentration, $\mu\text{mol/L}$ | 52.2 (13.3) | 52.2 (14.5) | 51.5 (11.0) | 53.1 (15.0) |
| Total iron binding concentration, $\mu\text{mol/L}$ | 65.0 [56.6;69.8] | 67.3 [56.6;70.7] | 62.9 [56.8;68.8] | 65.2 [56.0;69.2] |
| Transferrin saturation, % | 16.1 [9.00;27.4] | 19.4 [7.60;30.2] | 16.6 [11.0;22.8] | 13.2 [8.70;22.0] |
| Ferritin, $\mu\text{g/L}$ | 9.00 [6.00;15.5] | 7.50 [6.00;10.8] | 8.00 [7.00;12.2] | 13.0 [6.00;20.8] |
| White cell count, $10^9/\text{L}$ | 5.26 [4.34;6.67] | 5.73 [4.50;6.66] | 5.27 [4.37;6.74] | 4.92 [3.96;6.53] |
| Red cell count, $10^{12}/\text{L}$ | 4.41 (0.36) | 4.41 (0.34) | 4.41 (0.37) | 4.42 (0.40) |
| Hb, g/dL | 12.3 (1.25) | 12.0 (1.44) | 12.3 (1.27) | 12.4 (1.07) |
| Haematocrit, L/L | 0.38 (0.03) | 0.38 (0.03) | 0.38 (0.03) | 0.38 (0.03) |
| Mean cell volume, fL | 87.6 [83.8;89.9] | 86.8 [79.9;89.9] | 87.0 [84.9;89.6] | 88.7 [86.3;91.8] |
| Mean cell Hb, pg | 28.2 [26.4;29.5] | 27.9 [25.5;29.3] | 27.8 [27.3;29.1] | 28.6 [27.8;30.3] |
| Mean cell Hb concentration, g/dL | 32.1 (1.32) | 32.0 (1.51) | 32.1 (1.15) | 32.3 (1.34) |
| Red cell distribution width, % | 13.8 [13.1;14.6] | 13.8 [13.1;15.4] | 14.1 [13.0;14.5] | 13.7 [13.1;14.6] |
| Platelets, $10^9/\text{L}$ | 288 [229;334] | 296 [260;328] | 289 [204;333] | 264 [230;339] |
| Neutrophils, $10^9/\text{L}$ | 2.87 [2.33;3.98] | 3.09 [2.39;3.73] | 2.94 [2.46;4.52] | 2.64 [2.09;3.86] |
| Lymphocytes, $10^9/\text{L}$ | 1.68 (0.48) | 1.91 (0.52) | 1.63 (0.39) | 1.51 (0.48) |
| Monocytes, $10^9/\text{L}$ | 0.42 [0.33;0.55] | 0.40 [0.31;0.49] | 0.42 [0.36;0.55] | 0.42 [0.33;0.56] |
| Eosinophils, $10^9/\text{L}$ | 0.11 [0.08;0.21] | 0.11 [0.06;0.31] | 0.10 [0.08;0.16] | 0.14 [0.09;0.23] |
| Basophils, $10^9/\text{L}$ | 0.02 [0.02;0.04] | 0.02 [0.02;0.04] | 0.02 [0.02;0.03] | 0.04 [0.02;0.05] |

**Table 3.**
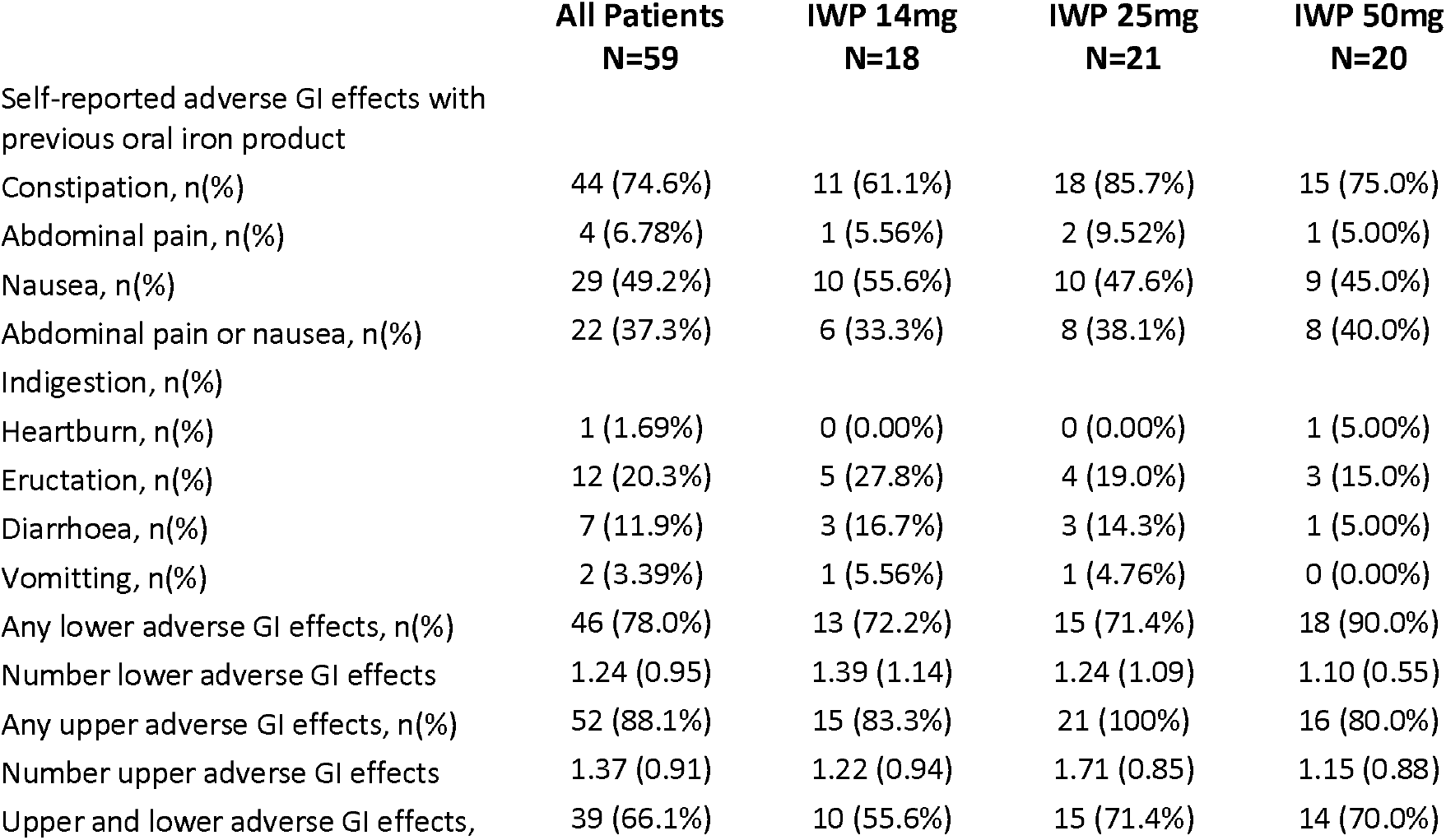
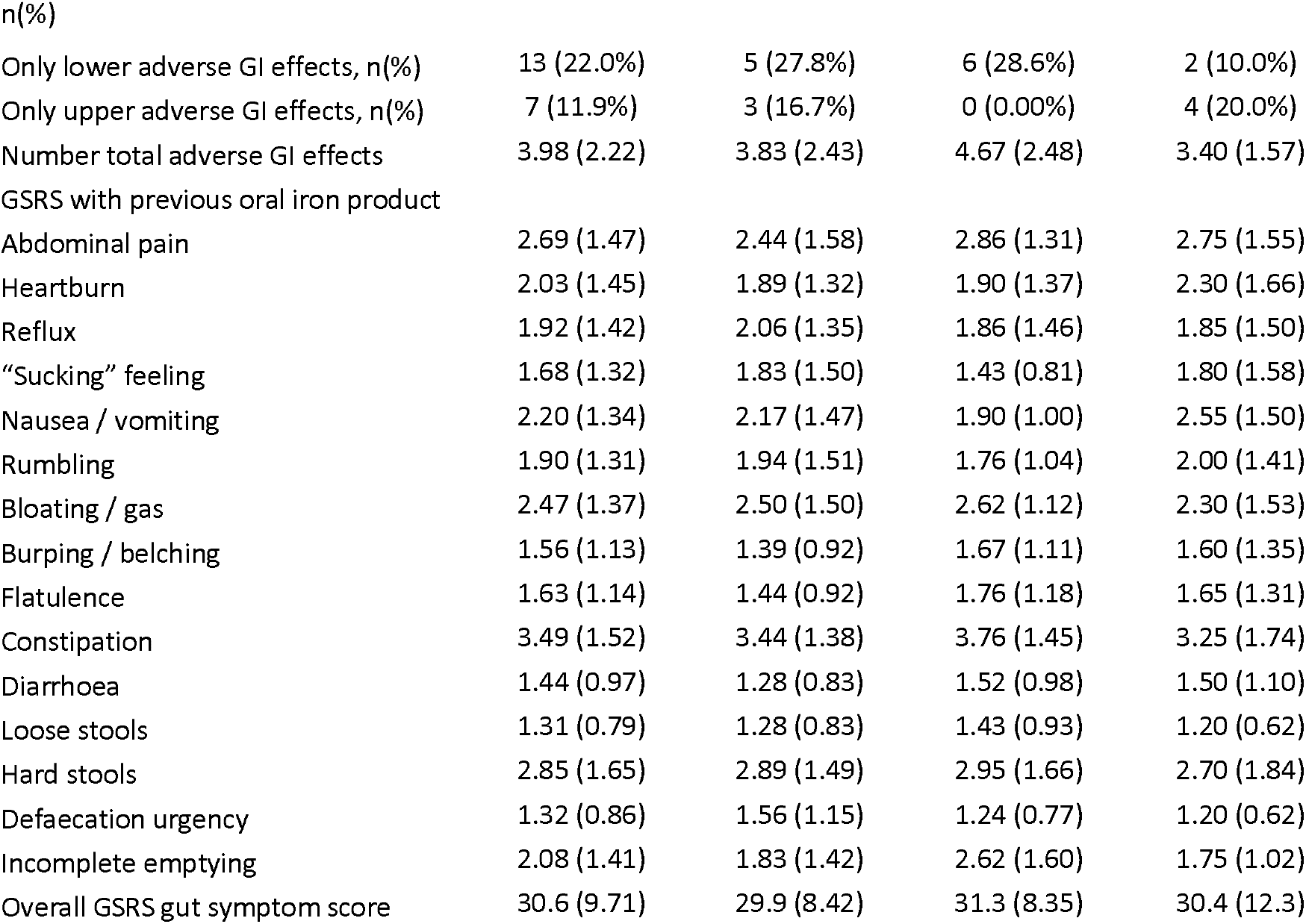
Self-reported gastrointestinal adverse effect profile and Gastrointestinal Symptom Rating Score (GSRS) associated with previous oral iron products of participants with ferritin <30µg/L and self-reported gastrointestinal intolerance to oral iron who were randomised to three different daily elemental iron doses of IWP (14mg, 25mg, 50mg). An overall GSRS gut symptom score of 15 is a perfect GSRS score reflecting no adverse GI symptoms. Abbreviations: IWP = iron-whey-protein formulation; GI = gastrointestinal; GSRS = Gastrointestinal Symptom Rating Scale gut symptoms score.

Following a minimum washout period of on average 9.8 ± 3.9 days, the overall GSRS score was significantly reduced in the cohort at the prospective study baseline (4.4 ± 7.05, Supplemental File Table S3) compared with the previous iron product (15.6 ± 9.71, p<0.001 vs baseline). There was a strong correlation between GSRS at baseline (following washout) and the GSRS reported with the previous iron product (OR 2.57, 95% CI 1.94-3.41, p<0.001) as well as the number of self-reported adverse GI effects on the previous oral iron product, (OR 3.03, 95% CI 1.33-6.92, p=0.009). This suggests the overall experience of adverse GI effects while taking prior oral iron could be related to the underlying adverse GI symptoms experienced by women when they are not taking oral iron.

### Adherence/persistence

The impact of IWP on overall adherence/persistence is presented in Figure 1(A). A total of 48 (81.4%) of the total cohort were classified as adherent/persistent with therapy using IWP compared to 12 (20.3%) taking the prior oral iron p<0.0001. Patients taking IWP were 4.0 (95% CI 2.4 to 6.7) times more likely to be adherent/persistent with IWP than with the previous oral iron (p<0.0001). Similar results were seen within the three dose groups and are presented in Supplemental File Figure S1, with 16 (88.9%), 17 (80.1%) and 15 (75.0%) participants taking 14mg, 25mg and 50mg respectively who were adherent/persistent with therapy. These were significantly higher than 4 (22.2%), 5 (23.8%) and 3 (15.0%) who persisted taking the previous oral iron in the respective 14mg, 25mg and 50mg groups (all p<0.001 versus IWP). The relative improvement in adherence/persistence was consistent across the three dose groups: 4.0 (95% CI 1.7 to 9.6) for IWP 14mg; 3.4 (95% CI 1.5 to 7.5) for IWP 25mg; 5.0 (95% CI 1.7 to 14.6) for IWP 50mg.

**Figure 1.**
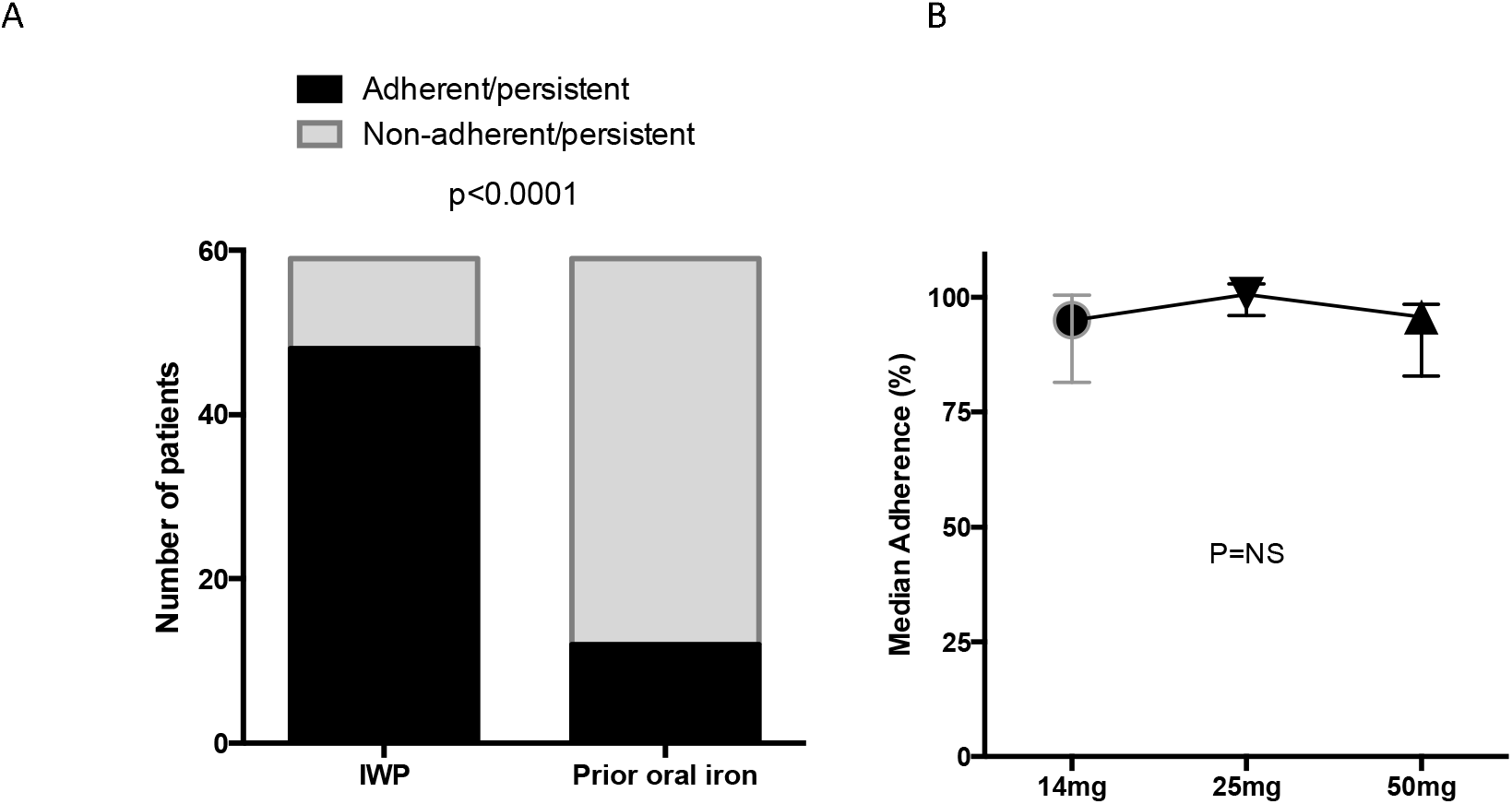
Overall adherence/persistence with IWP amongst 59 women with a history of intolerance to oral iron and ferritin < 30 ug/L compared to the previous oral iron (A). Also presented (B) is the median adherence using per dose group taking IWP over the study period. Abbreviation: IWP= iron-whey-protein formulation.

Median adherence with IWP was 96.4 (IQR 83.6%, 100.6%) over the course of 12 weeks and did not differ across the three dose groups (Figure 1B). This includes an adherence score of 0 attributed to 5 women who withdrew from the study following randomisation. Three of these women withdrew due to adverse GI symptoms which they reported as possibly, but not probably, due to IWP. One woman withdrew for personal reasons related to menorrhagia and one woman was lost to follow up. All 5 women were amongst 47 who had previously stopped taking oral iron due to adverse GI effects. Of the remaining 54 women who persisted with therapy, 48 demonstrated good average adherence (>80% based on pill counts)

### Gastrointestinal Symptom Rating Scale

In accordance with these data, the average GSRS score did not change from baseline at 6 weeks and 12 weeks post randomisation for the entire cohort (Figure 2A) and these scores were both significantly lower than the GSRS score reported with the previous oral iron product. Nor did the GSRS score change by dose or over the study period using repeated-measures-mixed-model analyses with adjustment for age, BMI and baseline GSRS. Overall, using reliable change index with 95% confidence,[18] 45 (81.8%) women had an improvement in GSRS using IWP compared to the previous iron product. Using reliable change index with 95% confidence, more women who previously took higher dose oral iron (>60mg elemental iron, n=35, 83.3%) had improved gut-symptom-scores on IWP compared versus those taking lower dose oral iron previously (n=10, 58.8%, P=0.045). Accounting for three further women who withdrew due to adverse GI effects and one woman lost to follow up, these data show 4.25 (95%CI 2.15 to 8.39, P<0.0001) more patients had adverse GI effects when taking the previous oral iron product. There was no difference between the three dose groups in terms of GSRS over the 12 weeks (Figure 2B).

**Figure 2.**
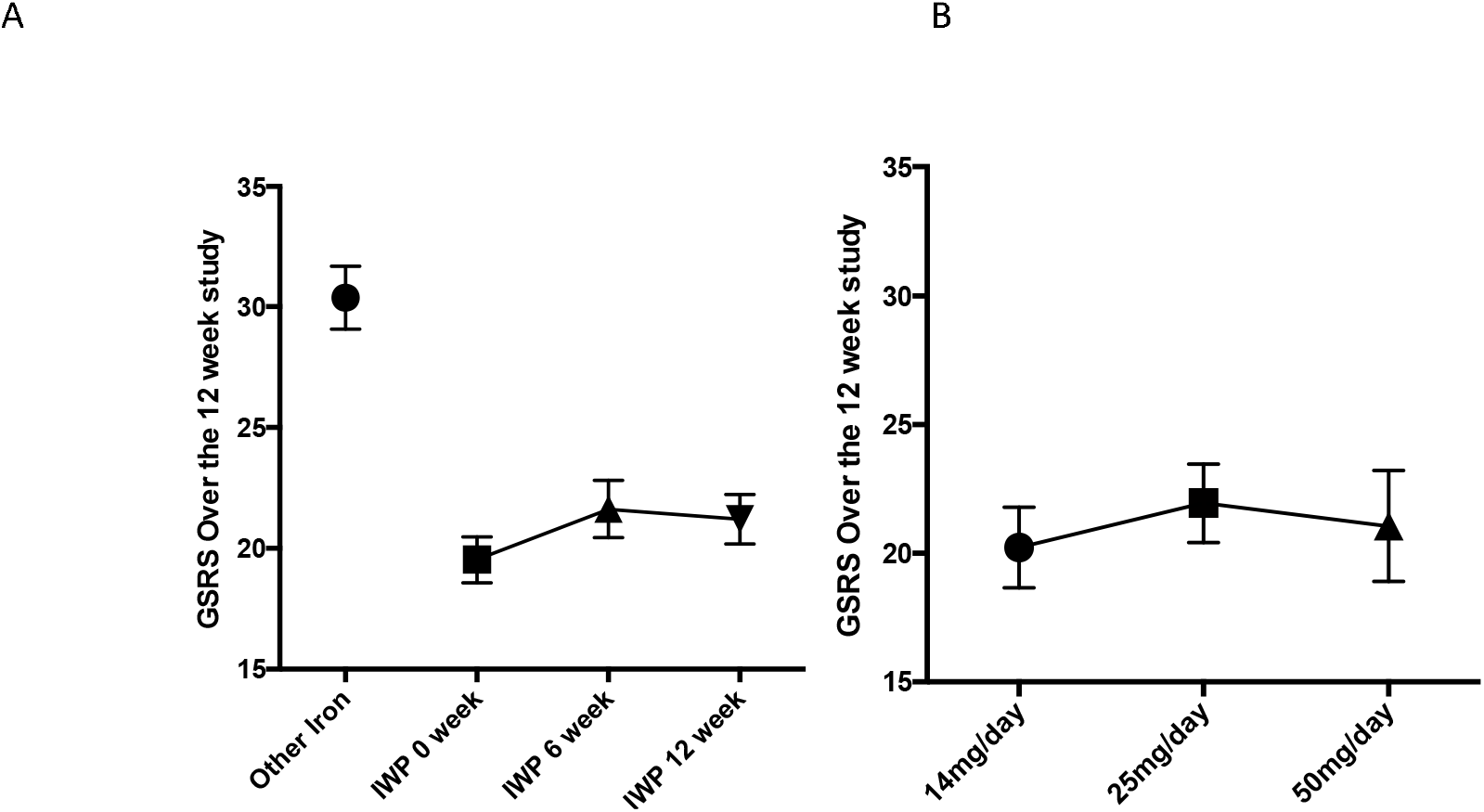
The overall Gastrointestinal Symptom Rating Score (GSRS) for previous oral iron product (black circle) and for IWP over the course of the study (A, all P<0.001 versus previous oral iron product) and (B) the average GSRS data by dose group (ANOVA P=NS) amongst 59 women with a history of intolerance to oral iron. An overall GSRS gut symptom score of 15 is a perfect GSRS score reflecting no adverse GI symptoms. Abbreviations: IWP= iron-whey-protein formulation; GSRS=Gastrointestinal Symptom Rating Scale gut symptoms score.

### Elicited Adverse Gastrointestinal Effects

All participants in the study had adverse GI effects that were probably associated with the previous oral iron product and on average 3.98 ± 2.22 adverse GI effects were attributed to the prior oral iron product. During the course of the prospective study, participants were questioned on four separate occasions about adverse GI effects, their severity and an assessment was made on whether they were possibly or probably associated with IWP. In total, participants reported six times fewer adverse events that were possibly or probably associated with IWP (0.59 ± 0.91, P<0.00001 versus prior oral iron product, Supplemental File Figure S2). Only one patient (4.3%) reported adverse GI effects (constipation, dark stools and excess flatulence) that were probably related to IWP. In this case, the patient was taking IWP 14mg, all symptoms were considered mild and the patient was happy to persist with treatment for 12 weeks. Coupled with the 5 people who withdrew from the study, at least 53 (89.9%) patients taking IWP were free of adverse GI effects and patients were at least 9.8 (95%CI 4.6 to 21.0, P<0.0001) times more likely to report adverse GI effects that were probably related to oral iron with the previous oral iron product. In addition, 22 participants reported side effects that were “possibly” related to IWP. Most of these (n=17, 77.3%) reported mild symptoms, 3 (13.6%) reported moderate symptoms and 2 patients (9.1%) reported severe symptoms.

Overall, patients were 4.0 (95%CI 2.3 to 7.0, P<0.0001) more likely to experience constipation and 3.2 (95%CI 1.7 to 6.2, P=0.0002) more likely to experience abdominal pain with the prior oral iron product than with IWP. A total of 44 (74.6%) and 29 (49.2%) patients had reported constipation and abdominal pain respectively attributed to the previous oral iron product. This reduced to 11 (18.6%) and 9 (15.3%) respectively with IWP. Four patients reported diarrhoea with the previous oral iron product compared with 2 patients taking IWP [relative risk 2.0 (95%CO 0.4 to 10.5, P=NS)].

### Effects on Ferritin, Transferrin Saturation and Haemoglobin

The analysis of serum iron, ferritin, transferrin saturation and haemoglobin in the overall cohort and by IWP dose group is presented in Table 4.

**Table 4.** Serum iron, ferritin, transferrin saturation and haemoglobin data in the overall cohort and in the three dose groups. Abbreviations: IWP= iron-whey-protein formulation; 6w=6 weeks; 12w=12 weeks; TSAT=transferrin saturation; Hb=haemoglobin.

|  | All<br>Participants<br>N=40 | IWP 14mg<br>N=12 | IWP 25mg<br>N=14 | IWP 50mg<br>N=14 | P<br>Value |
| --- | --- | --- | --- | --- | --- |
| Serum Iron baseline, $\mu\text{mol/L}$ | 11.3 (7.45) | 12.5 (7.42) | 11.2 (6.63) | 10.3 (8.58) | 0.774 |
| Serum Iron 6w, $\mu\text{mol/L}$ | 17.4 (9.10) | 18.3 (12.9) | 17.8 (7.91) | 16.1 (6.56) | 0.814 |
| Serum Iron 12w, $\mu\text{mol/L}$ | 20.5 (11.0) | 22.4 (13.1) | 20.6 (11.0) | 18.7 (9.52) | 0.700 |
| Serum iron change 6w, $\mu\text{mol/L}$ | 6.07 (9.43) | 5.83 (9.51) | 6.59 (9.68) | 5.76 (9.79) | 0.970 |
| Serum iron change 12w, $\mu\text{mol/L}$ | 9.19 (12.4) | 9.95 (13.2) | 9.36 (13.0) | 8.36 (12.0) | 0.949 |
| Ferritin baseline, $\mu\text{g/L}$ | 8.00 [6.00;13.0] | 8.00 [5.00;10.2] | 8.00 [7.00;12.2] | 9.00 [5.25;17.5] | 0.862 |
| Ferritin 6w, $\mu\text{g/L}$ | 17.0 [10.8;22.0] | 10.5 [7.00;15.8] | 17.5 [13.8;21.8] | 20.0 [12.5;24.2] | 0.037 |
| Ferritin 12w, $\mu\text{g/L}$ | 15.5 [9.00;24.2] | 8.50 [6.50;16.2] | 16.0 [11.2;22.5] | 20.0 [12.2;30.0] | 0.013 |
| Ferritin change 6w, $\mu\text{g/L}$ | 7.85 (10.1) | 2.83 (4.73) | 8.21 (6.65) | 11.8 (14.3) | 0.075 |
| Ferritin change 12w, $\mu\text{g/L}$ | 5.97 (8.02) | 1.42 (5.07) | 6.57 (7.09) | 9.29 (9.46) | 0.037 |
| TSAT baseline, % | 18.4 (13.4) | 20.6 (12.9) | 17.7 (11.8) | 17.2 (15.8) | 0.795 |
| TSAT 6w, % | 28.5 (13.8) | 28.8 (17.4) | 30.5 (14.1) | 26.3 (10.2) | 0.726 |
| TSAT 12w, % | 33.6 (17.6) | 35.1 (19.3) | 34.8 (19.5) | 31.2 (14.9) | 0.829 |
| TSAT change 6w, % | 10.1 (15.4) | 8.19 (13.5) | 12.8 (17.7) | 9.02 (15.3) | 0.722 |
| TSAT change 12w, % | 15.2 (19.5) | 14.4 (18.9) | 17.1 (22.0) | 14.0 (18.8) | 0.911 |
| Hb baseline, g/dL | 12.1 (1.17) | 11.9 (1.42) | 12.3 (1.06) | 12.2 (1.10) | 0.731 |
| Hb 6w, g/dL | 12.5 (0.93) | 12.3 (1.16) | 12.4 (0.93) | 12.8 (0.69) | 0.415 |
| Hb 12w, g/dL | 12.9 (0.95) | 12.7 (1.07) | 12.8 (0.97) | 13.2 (0.83) | 0.438 |
| Hb change 6w, g/dL | 0.30 [-0.02;0.82] | 0.55 [-0.12;0.90] | 0.25 [-0.10;0.68] | 0.35 [0.10;1.15] | 0.514 |
| Hb change 12w, g/dL | 0.65 [0.10;1.35] | 0.50 [0.18;1.25] | 0.60 [0.10;1.10] | 0.85 [0.32;1.65] | 0.550 |

Median ferritin levels overall increased from 8.00 [IQR 6.00;13.0] to 15.5 [IQR 9.00;24.2] µg/L at 12 weeks over the study (P=0.0002, Figure 2A). In addition, the mean changes within groups showed distinct differences according to the dose of IWP (P=0.035) with significant within-group increases in the 25mg and 50mg dose groups (Figure 2B). The mean ferritin increase within the 25mg dose group was 6.6 (95%CI 2.5 to 10.7) µg/L and within the 50mg dose group was 9.3 (95%CI 3.8 to 14.8) µg/L. The change in the 14mg dose group (1.6 (95%CI −1.4 to 4.6) µg/L did not reach statistical significance.

We pre-specified subgroup analyses of those patients with iron deficiency anaemia, where haemoglobin levels increased from 11.36 (95%CI 10.95 to 11.77) to 12.40 (95%CI 12.03 to 12.76, P=0.0007, Figure 3A). In addition, although there was no significant difference noted in the changes across each dose group using ANOVA, Figure 3B shows the mean changes at 12 weeks within each dose group (B) were significant in the 25mg and 50mg dose group only. The increase over time in patients with iron deficiency anaemia treated with IWP 50mg daily was 1.35 g/dL (95%Ci 0.54 to 2.16, P<0.01, Supplemental File Figure S3).

**Figure 3.**
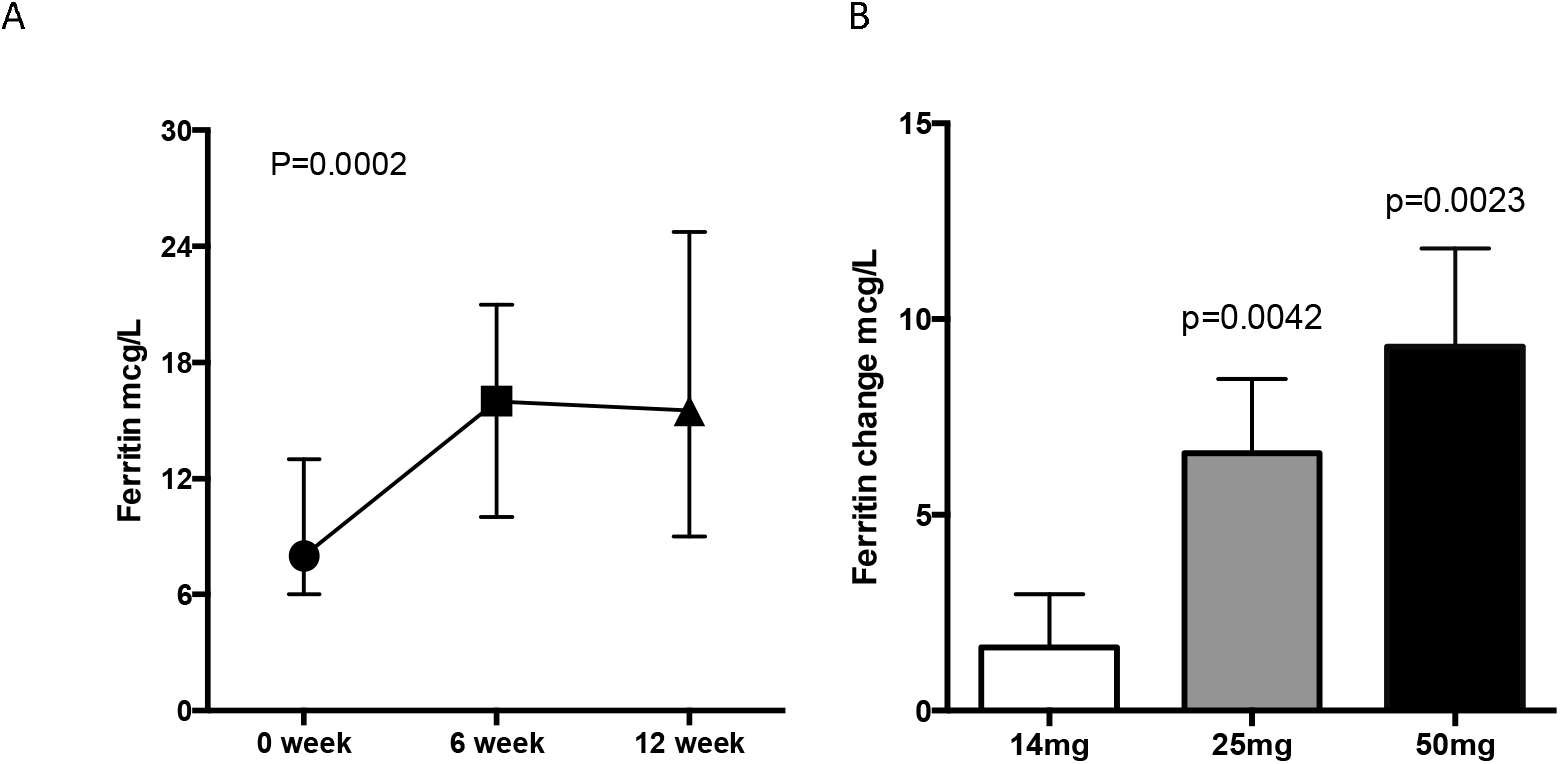
Median ferritin levels over time in the entire cohort (A) and mean changes from baseline to 12 weeks in each IWP dose group (B) amongst 59 women with a history of intolerance to oral iron and iron deficiency. Abbreviations: IWP= iron-whey-protein formulation.

In the total cohort (including those without anaemia), haemoglobin increased from 12.30 ± 1.25 to 12.89 ± 0.95 (p<0.001). The within-group increases were 0.60 (95%CI 0.08 to 1.1, P=0.023) g/dL in the 14mg dose group, 0.51 (95%CI 0.04 to 0.97, P=0.036) g/dL in the 25mg dose group and 0.96 (95%CI 0.22 to 1.71, P=0.01).

### Impact on Quality of Life

Detailed SF-36 data broken down according to the 8-health related quality of life domains are presented in Supplemental File Figure S4. Using ANOVA, there were significant differences across the domains (P<0.0001) and, as expected, the SF-36 Energy/Fatigue domain scores in this population (60 ± 4%) were significantly impaired compared to all the other domain scores at baseline (all P<0.001). Remarkably, there was no difference in the baseline SF-36 Energy/Fatigue domain scores between those with low iron stores (ferritin 12 µg/L – 30 µg/L, SF-36 Energy/Fatigue 61.1 ± 5.1%) and those with iron deficiency (ferritin <12 µg/L, SF-36 Energy/Fatigue 60.9 ± 4.5%). Furthermore, despite the presence of the pandemic, the Energy/Fatigue domain scores increased significantly in the overall group over the study period from 60.9 ± 3.4 % to 71.2 ± 2.6 % (P=0.0007) and significant within group changes were observed in the 25mg and 50mg daily dose groups. Interpretation of these data may be limited by the onset of Covid-19 pandemic throughout 2020 and, accordingly, they show a dramatic drop in the social (92 ± 2% to 79 ± 3%, p<0.0001) and emotional limitation 93 ± 2% to 87 ± 3%, P=0.035) domain scores. In a post-hoc analysis, the overall SF-36 score increased from 82.8 ± 1.7 to 85.6 ± 1.4, P=0.049 when these measures were excluded.

**Figure 3.**
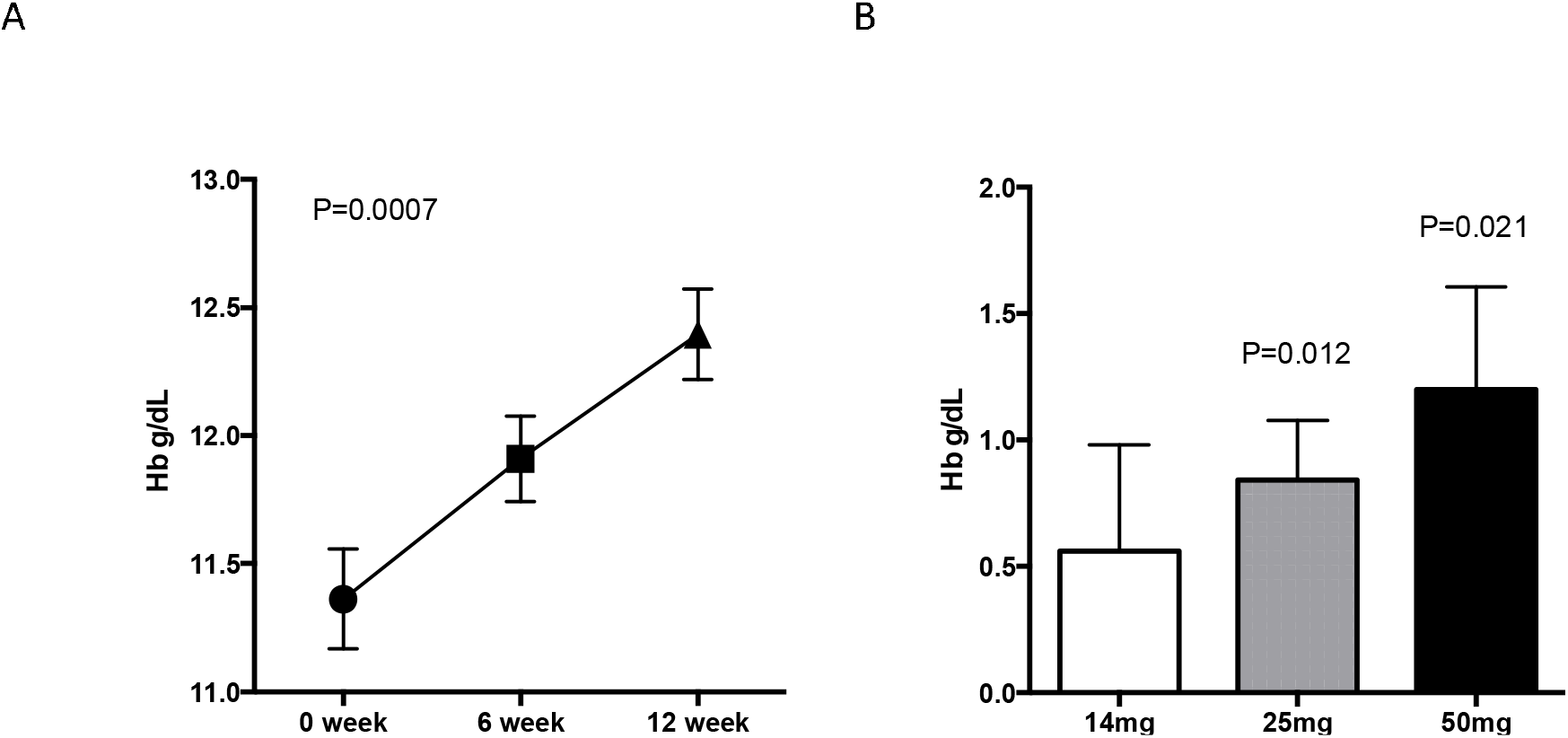
Mean haemoglobin levels over time (A) and mean changes from baseline to 12 weeks in each IWP dose group (B) in women with iron deficiency anaemia with a history of intolerance to oral iron. Abbreviations: IWP= iron-whey-protein formulation; Hb=haemoglobin.

**Figure 4.**
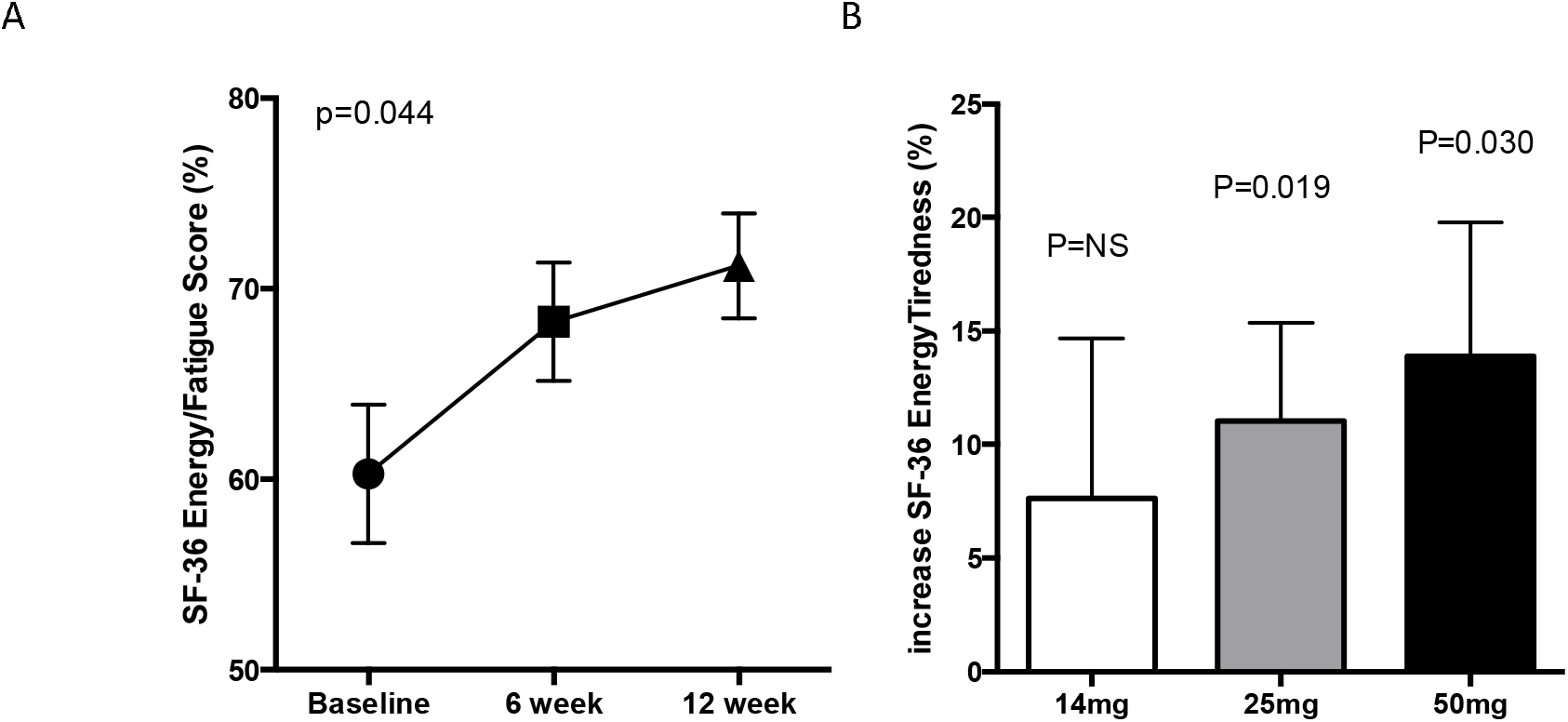
Mean SF-36 Energy/Fatigue Scores over time (A) and mean changes from baseline to 12 weeks in each IWP dose group (B) in women with iron deficiency anaemia with a history of intolerance to oral iron. Abbreviations: IWP= iron-whey-protein formulation; SF-36 = short form 36 health related quality of life score.

## DISCUSSION

Iron deficiency and iron deficiency anaemia are chronic and highly prevalent causes of morbidity that appear to be underdiagnosed in women of childbearing age.[1,2,19] Their management in the community relies on administration of oral iron products that are often poorly effective and/or cause intestinal side effects that limit adherence, persistence and may indeed contribute to continuation of the nutritional deficit.[3-5] In this study we have recruited women with low iron and iron deficiency with a history of intolerance to oral iron to investigate the extent of this problem as well as a potential management approach. A majority (62%) of the women in the present screening study had low iron stores, over 3 in 10 had moderate to severe iron deficiency and 1 in 6 had iron deficiency anaemia. These results are in accordance with findings on iron deficiency and iron deficiency erythropoiesis in adult women who are frequent blood doners.[19] The adverse GI effect most commonly reported in this group was constipation (79%) and a majority (58%) of women also reported combined upper and lower GI adverse effects. The causes of GI adverse effects in oral iron use remain poorly understood. Endoscopic reports show direct damage from iron deposition in the upper GI tract, which may have a contribution from iron redox activity and associated reactive oxygen species (ROS) generation.[7,8,20] Pathophysiological shifts in microbiota composition may contribute to lower intestinal issues, especially at high dose.[9,20] Oral iron absorption, which is hepcidin regulated, is also affected by iron dose but also by intestinal inflammation associated with oral iron intolerance. [3,5,21] Oral iron treatment of iron deficiency in an iron intolerant group is for these reasons highly challenging.

From the group of pre-menopausal adult women, a subgroup that were iron deficient or iron deficient and anaemic were selected for a prospective trial to investigate a potential management approach. IWP has been reported to be well absorbed and to produce relatively little ROS in intestinal cell models.[17] Iron deficiency and iron deficiency anaemia has been conventionally treated with oral elemental iron doses in the range 100-200 mg daily. [22] There has been a shift in treatment guidance to lower doses in the past decade with recommendations towards the range 60-100 mg daily.[3,10,23] Considering its reported high bioavailability, we have investigated IWP in the dosing range 14-50 mg daily in the randomised prospective part of this study. Using the validated GSRS gut-symptom-score to systematically track adverse GI effects for the first time, treatment with IWP resulted in better gut-symptom-score, six times fewer elicited adverse GI events and four times better compliance than patients reported experience with prior oral iron.

There was no difference between the three IWP dose groups (14mg, 25mg, 50mg daily elemental iron) in terms of compliance or tolerability, suggesting higher IWP doses can be used, particularly in those with iron deficiency anaemia. Finally, these patients had impaired ferritin, transferrin saturation and haemoglobin levels as well as reduced SF-36 energy and fatigue domain scores at baseline. Over a 12 week period, overall improvements in these parameters were observed, particularly in the 25mg and 50mg dose groups. Taken together, these data show that a self-report of oral iron intolerance in adult women of childbearing age is a harbinger of adverse clinical status related to low iron stores, which is modifiable using IWP.

More than 2.2 billion people worldwide have anaemia and half of this burden is caused by iron deficiency, while a further billion people are estimated to have iron deficiency without anaemia [1]. The present study underlines that while iron deficiency anaemia has been, and frequently remains, synonymous with iron deficiency, the latter is a broader condition that can occur early in the natural history of iron deficiency anaemia. Iron deficiency can affect other organs/tissues, such as hair growth, immune function, skeletal muscles and the heart, long before there is evidence of impaired erythropoiesis.[24,25] The commonest causes of iron deficiency are inadequate intake, poor absorption of iron [3,24] and blood loss, in particular due to menstruation [1,2,25]. Pre-menopausal adult women are at high-risk of iron deficiency and anaemia because of inadequate iron intake and menstrual blood loss.[1,2,11,24] While the estimates vary on the prevalence of iron deficiency amongst adult women in different geographic locations, between 10-20% of menstruating women in the UK have been shown to be iron deficient [1,24,25]. Our study may build on this literature by highlighting a group of women with higher rates of iron deficiency, iron deficiency anaemia and low iron stores. Although detailed information on the reasons for this relatively high prevalence of low iron stores were not collected in the screening study, baseline analysis of those included in the prospective study are instructive in this regard. Firstly, the majority of these women (77.8%) had to stop taking their oral iron because of adverse GI effects, principally constipation, abdominal pain and nausea. Second, 6 in 10 self-reported heavy menstrual periods. Third, there was a strong, multivariable correlation observed between GSRS gut-symptom-score while taking all oral iron products and the baseline score following at least one week of washout from the prior oral iron. This suggests that women with underlying adverse GI symptoms, including those with undiagnosed coeliac disease or irritable bowel syndrome, may risk more severe adverse GI effects with oral iron, exacerbating compliance and deficiency states [20, 26].

The prospective study also shows that few women with low iron stores had a prior diagnosis of iron deficiency or anaemia. This may arise from differences in diagnostic thresholds commonly used in practice and evaluation of ferritin levels only in those with pre-existing anaemia.[25, 28] In the absence of inflammation or infection, low serum ferritin levels are the hallmark of absolute iron deficiency, reflecting exhausted stores. While many hospital and commercial laboratories use a diagnostic threshold of 12 µg/L, which is highly specific, it is not adequately sensitive for the diagnosis of iron deficiency in pre-menopausal women. [27] Furthermore, absent bone marrow iron stores and impairment of iron erythropoiesis commonly occurs in women with serum ferritin levels in in the range 25 to 40 µg/L.[27] In our study, the 12 µg/L threshold is more commonly associated with iron deficiency anaemia, however it is increasingly recognised that ferritin levels of < 30 µg/L, termed “low iron stores” in our study and elsewhere [28], represent a form of mild absolute iron deficiency. The importance of this is reflected in our finding that iron deficiency doubles in prevalence to over 6 in 10 women with inclusion of mild disease, using a 30 µg/L threshold. Although fewer of these women had anaemia, the SF-36 data suggest that ferritin between 12 and 30 µg/L, is associated with significant impairment of daily energy and fatigue. Furthermore, there was no difference between energy and fatigue domain scored between those patients and those with moderate to severe iron deficiency (ferritin <12 µg/L). Nor was there a difference in treatment response.

Overall, these data underline the need to re-evaluate screening and treatment algorithms for iron deficiency as well as the “reactive” practice of evaluating iron stores only in women with anaemia. The high prevalence of heavy menstrual periods in our study may explain advice received, or a decision to use oral iron products without a formal diagnosis of iron deficiency or anaemia. This also underlines that the commonest global cause of iron deficiency is menstruation [1,2,11,25,28] and that this deficiency is linked to greater tiredness and lower energy levels. These data may argue for improved nutritional self-care for women with periods as well as greater awareness of low iron or mild iron deficiency in the absence of anaemia amongst adult women of childbearing age.

The frequency and range of upper and lower adverse GI effects associated with oral iron in the screening study highlights challenges of GI adverse effects with oral iron and their association with poor compliance. [4,5,11] The majority of women in our study experienced constipation and/or abdominal pain, in accordance with a previous systematic review.[11] The high prevalence of constipation underlines the difficulty in advocating delayed release or enteric coated oral iron products as a solution to poor GI tolerability, as they have relatively poor absorption,[13] increasing the unabsorbed iron load reaching the bowel, therefore potentially aggravating constipation. Interestingly, all of the women who had taken enteric coated ferrous sulfate products in our study reported experiencing constipation. Although ferrous sulfate has been considered the gold standard oral iron [29], and is poorly tolerated [3-5], the majority of women in our prospective study were had been taking high-dose, immediate release ferrous fumarate and it is interesting to note that the improvement in tolerability with IWP over prior iron products was greater in patients previously taking higher dose oral iron (elemental iron dose >60mg).

This is the first trial to use the validated GSRS gut-symptom-score to prospectively track GI tolerability over time with a specific, new oral iron treatment. The data show no change overall in average GSRS score over time as well as significantly lower GSRS scores and at least 4 times better gut symptom scores with IWP versus the prior oral iron product. Furthermore, the GSRS data also accord with the observation that only 4 patients attributed adverse GI events to IWP across the prospective study, and there were six times fewer adverse GI events reported overall with IWP based on elicited adverse GI event data obtained at 4 timepoints during the 12 week study period. Three of these women stopped taking IWP due to adverse GI effects. Overall, in agreement with published *in-vitro* data showing significantly reduced iron oxidative stress in gut cells with IWP compared with ferrous sulfate, [17] the prospective study provides clinical evidence of low adverse GI effects as well as high adherence and persistence rates in this vulnerable cohort.

The data also show good tolerability and high adherence across the dose range of IWP studied, providing reassurance when using higher IWP doses to treat women with iron deficiency and a history of oral iron GI intolerance. There was no difference in GSRS data across different doses of IWP, although we did not note differences in GSRS score between those taking higher versus lower doses of the prior oral iron product. IWP has been shown to have improved bioavailability versus the WHO gold-standard ferrous sulfate [29], which has low fractional absorption [3]. Using 50 mg IWP daily in the subset with mild-to-moderate iron deficiency anaemia, haemoglobin significantly increased by 1.35 g/dL over 12 weeks and was normalised in most women. Although some women did respond to IWP at the nutritional reference value dose 14mg, the data do not support use of this dose in women with iron deficiency and it should be reserved as a supplement for maintenance of normal iron stores. Conversely, there is a consistency of beneficial response across ferritin, transferrin saturation, haemoglobin and clinical (SF-36 Energy/Fatigue) measures with the 25 mg and 50 mg daily dose groups. There are other pharmacotherapeutic ways to improve iron stores and anaemia amongst women with a history of intolerance to oral iron, particularly using infusions of iron-carbohydrate complexes.[6,25,28] However, guidelines continue to support the preferential use of oral formulations of iron first line and the present data support intervention using higher doses of IWP to maximise outcomes in women with mild-to-moderate iron deficiency with or without anaemia.

There are a number of limitations to this study. First, the purpose of the study was to understand the haematinic morbidity associated with self-reported intolerance to oral iron and its management with different doses of IWP. There is no direct comparison with other iron products nor any placebo control. Second, there is a reliance in the study on self-report of gastrointestinal intolerance and adherence, although we tried to mitigate this with use of validated GSRS gut symptom scores. Third, the population was selected on the basis of a previous negative experience of oral iron, which may have introduced selection bias. These data should be confirmed in separate studies in different settings (e.g. primary care, secondary care). Finally, the study was not powered on overall SF-36 scores, which were further subject to potential bias in certain domains because of Covid 19 lockdown restrictions. In addition, it was not possible to obtain follow up bloods in a large minority of patients due to Covid 19. Despite this, there was adequate power overall and within groups to adjudicate treatment effects on adherence, self-reported adverse GI effects, ferritin and haemoglobin.

## Conclusions

Low iron, iron deficiency and anaemia are common in women of childbearing age with a history of intolerance to oral iron. Few of the women reported a history of diagnosed iron deficiency or anaemia. The data underline growing awareness that low iron stores (e.g. ferritin between 12 µg/L and 30 µg/L) represent a form of mild iron deficiency and are associated with similar fatigue and energy impairment as moderate to severe iron deficiency (ferritin < 12 µg/L). Accordingly, iron stores should be evaluated and managed in symptomatic women of childbearing age independently of the presence of anaemia. Finally, these data concur with a growing literature that high doses of oral ferrous salts may not be needed to improve iron stores and haemoglobin. IWP can improve self-reported oral iron adherence and tolerability as well as iron stores, haemoglobin and tiredness in these women.

## Data Availability

Consent has not been obtained for publication of patient level data.

## Acknowledgements

The authors acknowledge the support of the Atlantia Clinical Trials, Contract Research Organisation responsible for conduct of the PRECISION study according to GCP.

## Abbreviations

BMI: Body mass index
Bpm: beats per minute
CI: Confidence interval
DBP: Diastolic blood pressure
GI: Gastrointestinal
GSRS: Gastrointestinal Symptom Rating Scale gut symptoms score
Hb: haemoglobin
HR: Heart rate
IQR: Interquartile range
IWP: iron-whey-protein formulation
OR: Odds Ratio
SBP: Systolic blood pressure
SD: standard deviation
TSAT: transferrin saturation

