## Supplemental File for "Iron deficiency in Women of Childbearing Age with Self-reported Oral Iron Gastrointestinal Intolerance and Management with an oral Iron-Whey-Protein Formulation"

PRospectivE study of women of Childbearing age with gastrointeStinal Intolerance to Oral Iron (**PRECISION**), a randomised, prospective, double-blind, double dummy clinical trial.

Gilmer JGF, PhD, Research Director, Head of School. [1,2]
Fiona Ryan, PhD Director of Clinical Trials. [2]
Anna Seoighe MSc, Research Pharmacist
Maria Jose Santos-Martinez, MD, Associate Professor, Clinical Medicine[1]
Mark Ledwidge, PhD, Research and Development Director, Adjunct UCD Professor. [1,2,3]

**Abstract**

**Background**

Intolerance to oral iron is thought to result in poor adherence and persistence of nutritional deficit amongst women of childbearing age, however few studies have evaluated oral iron intolerance, iron deficiency and anaemia in this setting. Iron-whey protein microspheres (IWP) could help.

**Methods**

We documented self-reported oral iron gastrointestinal intolerance, ferritin and haemoglobin levels in a screening study of women of childbearing age. Following a washout period of 16 days, we randomised 59 of these women with iron deficiency, stratified according to the presence of anaemia, to three doses of IWP: (14mg daily, 25mg daily and 50mg daily). We excluded those with established gastrointestinal disease, potential allergy to whey protein and severe anaemia. The primary endpoint was persistence and adherence (>80% based on pill-counts). Secondary endpoints included changes in self-reported oral iron gastrointestinal intolerance, gastro-intestinal symptom rating scale (GSRS), serum iron, serum ferritin, transferrin saturation and haemoglobin levels.

**Results**

A total of 128 (62.7%) of the participants had low iron stores (ferritin < 30 µg/L), 65 (31.9%) had moderate to severe iron deficiency (ferritin <12 µg/L) and 33 (16.2%) had iron deficiency anaemia. Amongst 59 women who participated in the prospective study, 48 (81.4%) were classified as adherent/persistent with therapy using IWP compared to 12 (20.3%) taking the prior oral iron p<0.0001. These patients also showed significantly fewer reports of gastrointestinal intolerance with IWP ($0.59 \pm 0.91$) and lower GSRS scores ($6.2 \pm 7.5$) compared to the previous oral iron product ($3.98 \pm 2.22$, and $15.6 \pm 9.7$ respectively, both P<0.0001). There were no differences in adherence, self-reported adverse GI effects and GSRS between the dose groups during the study. Serum iron levels increased across the whole cohort from 11.3 ± 7.4 µmol/L to 20.5± 11.0 µmol/L (P<0.0001), transferrin saturation levels increased from 18.4 ±13.3 % to 33.6 ±17.6 % (P<0.0001) and median ferritin levels overall increased from 8.00 [IQR 6.00;13.0] to 15.5 [IQR 9.00;24.2] µg/L at 12 weeks (P=0.0002). Haemoglobin levels increased from 11.36 g/dL (95%CI 10.95 to 11.77) to 12.40 g/dL (95%CI 12.03 to 12.76, P=0.0007) in patients with anaemia and were normalised in most patients taking 50mg IWP daily.

**Conclusions**

Low iron, iron deficiency and anaemia are common in women of childbearing age with a history of intolerance to oral iron. Patients with low iron (ferritin < 30 µg/L) and moderate to severe iron deficiency (ferritin <12 µg/L) have similar impairment of energy. IWP can improve self-reported oral iron adherence and tolerability as well as iron stores, haemoglobin and tiredness in these women.

**Detailed study procedure**

**Screening cohort study.**

Subjects were recruited through adverts in local newspapers, online parenting website (www.rollercoaster.ie), general practitioners offices and the Atlantia Clinical Research Organisation patient database. Subjects underwent an initial phone screen to confirm their demographics, medical history (including absence of diagnosed gastrointestinal disease) and definite history of gastrointestinal intolerance to oral iron. Consenting, eligible subjects were then asked to omit their oral iron product for at least 7 days and were invited to a study screening visit in the clinic to investigate iron stores, haemoglobin, vital signs (blood pressure, heart rate & temperature), anthropometric measurements (weight, height & body mass index), medical history, history of iron deficiency, history of anaemia and self-reported history of intolerance to oral iron products. The self-reported history collected detailed measures of upper GI symptoms (e.g. self-reported history of stomach pain, nausea, vomiting, eructation, heartburn, indigestion) and lower GI symptoms (e.g. bowel habit changes, diarrhoea, constipation, stool discolouration). A fasting blood sample (16mls) was collected and ferritin and haemoglobin was measured.

**Prospective treatment study.**

The prospective trial evaluated the adherence, tolerability and efficacy of IWP overall and of three different elemental iron doses of IWP in women with a history of intolerance to oral iron, low iron or iron deficiency, with or without anaemia. This was carried out during three clinic visits (baseline, week 6 and week 12) and two phone call visits (weeks 3 and 9). In addition, there was stratified randomisation to include up to 30 patients with iron deficiency and mild to moderate anaemia as well as up to 30 patients with iron deficiency without anaemia.

Eligible women between 18 and 55 years of age, with a self-reported history of intolerance to oral iron and low iron stores (ferritin < 30 µg/L) or iron deficiency (ferritin < 12 µg/L) with or without mild to moderate anaemia (haemoglobin ≥9.5 g/dL and <12.0 g/dL) were invited to participate in the prospective treatment study. Once the screening blood results were reviewed and eligibility for the prospective study was confirmed following a washout period of at least one week from the previous iron product, the subject returned to the clinic for full clinical assessment. At visit 2 (baseline of the prospective study), a fasting blood sample was collected for evaluation of ferritin, serum iron, unbound iron binding capacity, transferrin saturation, and full blood count (including haemoglobin). A urine sample was collected, and pregnancy test performed. Subjects were queried about any changes in their health status and any non-treatment emergent events since the screening visit and medications were recorded. This included details of the prior oral iron product taken (dose, form, dosage frequency). Self-reported adherence, persistence, upper adverse GI effects (stomach pain, nausea, vomiting, eructation, heartburn, indigestion) and lower adverse GI effects (bowel habit changes, diarrhoea, constipation, stool discolouration) with the prior oral iron product were documented. In addition, a record was made of intolerance with the prior oral iron product was made using the Gastrointestinal Symptom Rating Scale (GSRS, details in Reference [14]). Subjects completed the following questionnaires: baseline GSRS reflecting GI symptoms at least one week after stopping the prior oral iron product, Short Form-36 (SF-36) health related quality of life (https://www.rand.org/health-care/surveys_tools/mos/36-item-short-form.html) and a menstrual period questionnaire (details in Supplemental File) including the date, duration and blood flow of their last menstrual period.

Subjects were randomized into one of three treatment groups, but were blinded as to which group they were in using double-blind, double-dummy masking. Subjects were supplied with a six-week supply of study product and instructions of dosing. Subjects were instructed to follow their habitual diet and exercise routine and to not consume any disallowed medications/supplements (e.g. supplements that can chelate iron) that could interfere with the assessment of the study product for the duration of the study. Subjects were provided with an appointment to return to the study site at week 6.

At week 3 and 9, during phone visits, all subjects were asked to complete the GSRS questionnaire, and the menstrual period questionnaire, to record the date, duration and blood flow of their last menstrual period. They also were queried about adverse events and relatedness with IWP (unrelated, possibly-related, probably-related). Subjects returned to the clinic site at week 6 and 12. At each clinic visit, any changes in health status, medications and any adverse events, including relatedness with IWP, were documented. A fasting blood sample was collected and a full blood count, including ferritin, serum iron, transferrin saturation was measured. Subjects completed the following questionnaires: GSRS, the Short Form-36 (SF-36) and the date, duration and blood flow of their last menstrual period using the Menstrual Period Questionnaire. Subjects returned any unused study product and additional product was dispensed at clinic visit 3 (week 6). The adherence based on pill-count was determined at weeks 6 and 12. Subjects were instructed to continue following their habitual diet and exercise routine and to not consume any disallowed medications/supplements that could interfere with the assessment of the study product for the duration of the study.

**Outcome measures and statistical analyses.**

The outcome measures in the screening study were prevalence of low-iron (ferritin <30 ug/L), moderate to severe iron deficiency (ferritin <12 ug/L) and anaemia (haemoglobin <12 g/dL) as well as the number and profile of self-reported adverse GI effects with oral iron. The primary outcome measure in the prospective study was the change in proportion of subjects adherent and persistent with therapy over the study period. Secondary outcomes evaluated from baseline to 12 weeks were: the difference in self-reported incidence of upper GI symptoms; the difference in self-reported incidence of lower GI symptoms; the change in gastrointestinal tolerability using GSRS; the change in haemoglobin in those with anaemia; the change in ferritin; the change in transferrin saturation; the change in tiredness and energy HRQOL using SF36.

**Menstrual Period Questionnaire**

MENSTRUAL PERIOD QUESTIONNAIRE

Do you suffer from heavy periods?

☐ Yes ☐ No

How long does your menstrual period usually last?

☐ less than 3 days

☐ 3 to 7 days

☐ more than 8 days

Do you experience menstrual flow that soaks through one or more pads or tampons every hour?

☐ Yes ☐ No

Do you need to use double sanitary protection to control your menstrual flow?

☐ Yes ☐ No

Do you experience menstrual flow that includes large blood clots?

☐ Yes ☐ No

Does your menstrual flow inferfere with your regular lifestyle?

☐ Yes ☐ No

Do you experience pain during your period?

☐ Yes ☐ No

Do you have other symptoms like easy bruising?

☐ Yes ☐ No

Do you take medications during your period?

☐ Yes ☐ No

If yes, please give details

______________________________________________________________________

______________________________________________________________________

______________________________________________________________________

____________________

**Complete each time you have your period:**

- Date of onset of menstruation (day/month/year):____________
- Days of menstruation: __________days
- Number of heavy blood loss days during menstruation: ________days

Indicate the number and type of pads and/or tampons used at heaviest blood loss day of menstruation, both during the day and night:

| | Number of pads | | | | Number of tampons | | | |
|---|---|---|---|---|---|---|---|---|
| | Mini/salvaslip | Normal | Super | Night / superplus | Mini/salvaslip | Normal | Super | Night / superplus |
| Day | | | | | | | | |
| Night | | | | | | | | |

### Additional Data

| | All Participants N=204 | Ferritin ≥ 12µg/L N=139 | Ferritin < 12µg/L N=65 | P value |
|---|---|---|---|---|
| Age, years | 36.6 (10.1) | 36.3 (9.87) | 37.3 (10.6) | 0.527 |
| History of iron deficiency, n(%) | 17 (8.33%) | 6 (4.32%) | 11 (16.9%) | 0.006 |
| History of anaemia, n(%) | 23 (11.3%) | 10 (7.19%) | 13 (20.0%) | 0.011 |
| History of iron deficiency or anaemia, n(%) | 26 (12.7%) | 13 (9.35%) | 13 (20.0%) | 0.042 |
| Weight, kg | 70.7 [60.7;81.4] | 70.6 [59.9;81.9] | 73.0 [63.2;79.6] | 0.909 |
| Height, m | 1.65 (0.06) | 1.65 (0.06) | 1.65 (0.06) | 0.900 |
| BMI, kg/m$^2$ | 25.8 [22.3;29.6] | 25.6 [22.3;29.8] | 26.2 [22.6;29.4] | 0.837 |
| Temp, °C | 36.3 (0.47) | 36.3 (0.47) | 36.1 (0.44) | 0.003 |
| SBP, mmHg | 109 [102;117] | 109 [102;117] | 108 [101;118] | 0.341 |
| DBP, mmHg | 73.8 (9.38) | 74.3 (9.38) | 72.9 (9.39) | 0.340 |

| | | | | |
|---|---|---|---|---|
| HR, bpm | 70.0 [65.0;77.0] | 70.0 [65.0;78.0] | 69.0 [65.0;76.0] | 0.304 |
| Smoking status: | | | | 0.519 |
| Never smoked, n(%) | 133 (65.2%) | 89 (64.0%) | 44 (67.7%) | |
| Previous smoker, n(%) | 47 (23.0%) | 35 (25.2%) | 12 (18.5%) | |
| Current smoker, n(%) | 24 (11.8%) | 15 (10.8%) | 9 (13.8%) | |
| Alcohol consumption, units/week | 2.64 (2.96) | 2.64 (2.99) | 2.63 (2.92) | 0.985 |
| Depot contraceptive, n(%) | 8 (3.92%) | 7 (5.04%) | 1 (1.54%) | 0.440 |
| Patch or ring contraceptive, n(%) | 15 (7.35%) | 13 (9.35%) | 2 (3.08%) | 0.152 |
| Oral contraceptive, n(%) | 37 (18.1%) | 27 (19.4%) | 10 (15.4%) | 0.615 |
| Serum Iron, µmol/L | 14.7 [9.25;21.4] | 17.4 [13.6;23.1] | 8.00 [5.90;14.9] | <0.001 |
| Total iron binding concentration, µmol/L | 61.0 (9.94) | 57.2 (8.39) | 66.7 (9.41) | <0.001 |
| Transferrin saturation, % | 25.7 [13.7;35.5] | 30.8 [23.6;42.7] | 11.9 [8.00;23.3] | <0.001 |
| Ferritin, µg/L | 18.0 [9.00;43.2] | 33.0 [18.0;53.0] | 7.00 [5.00;9.00] | <0.001 |
| White cell count, $10^{-9}$/L | 5.60 [4.76;6.79] | 5.50 [4.82;7.02] | 5.69 [4.55;6.25] | 0.334 |
| Red cell count, $10^{-12}$/L | 4.42 [4.24;4.70] | 4.46 [4.23;4.81] | 4.39 [4.27;4.62] | 0.301 |
| Haemoglobin, g/dL | 13.0 [12.2;13.7] | 13.4 [12.7;14.0] | 12.2 [11.4;12.9] | <0.001 |
| Haematocrit, L/L | 0.40 [0.37;0.42] | 0.41 [0.39;0.42] | 0.37 [0.36;0.39] | <0.001 |
| Mean cell volume, fL | 88.8 [85.4;92.1] | 90.4 [87.7;93.6] | 86.8 [81.1;88.9] | <0.001 |
| Mean cell Hb, pg | 28.9 [27.3;30.4] | 29.6 [28.3;30.9] | 27.4 [25.4;29.4] | <0.001 |
| Mean cell Hb concentration, g/dL | 32.4 (1.19) | 32.7 (1.05) | 31.8 (1.21) | <0.001 |
| Red cell distribution width, % | 13.3 [12.8;14.2] | 13.0 [12.6;13.5] | 13.9 [13.2;15.2] | <0.001 |
| Platelets, $10^{-9}$/L | 287 [247;324] | 279 [244;320] | 292 [248;335] | 0.596 |
| Neutrophils, $10^{-9}$/L | 3.26 [2.49;4.21] | 3.30 [2.49;4.43] | 3.20 [2.49;4.02] | 0.329 |
| Lymphocytes, $10^{-9}$/L | 1.73 [1.36;2.06] | 1.76 [1.38;2.07] | 1.67 [1.31;1.86] | 0.237 |
| Monocytes, $10^{-9}$/L | 0.44 [0.35;0.53] | 0.41 [0.34;0.53] | 0.46 [0.38;0.53] | 0.251 |
| Eosinophils, $10^{-9}$/L | 0.14 [0.08;0.26] | 0.13 [0.08;0.23] | 0.16 [0.08;0.29] | 0.372 |
| Basophils, $10^{-9}$/L | 0.03 [0.02;0.04] | 0.03 [0.02;0.04] | 0.03 [0.02;0.04] | 0.771 |
| Constipation, n(%) | 163 (79.9%) | 114 (82.0%) | 49 (75.4%) | 0.361 |
| Abdominal pain, n(%) | 12 (5.88%) | 9 (6.47%) | 3 (4.62%) | 0.756 |
| Nausea, n(%) | 118 (57.8%) | 76 (54.7%) | 42 (64.6%) | 0.235 |
| Abdominal pain or nausea, n(%) | 81 (39.7%) | 52 (37.4%) | 29 (44.6%) | 0.409 |
| Indigestion, n(%) | 60 (29.4%) | 35 (25.2%) | 25 (38.5%) | 0.076 |
| Heartburn, n(%) | 4 (1.96%) | 2 (1.44%) | 2 (3.08%) | 0.594 |
| Eructation, n(%) | 27 (13.2%) | 19 (13.7%) | 8 (12.3%) | 0.964 |
| Diarrhoea, n(%) | 24 (11.8%) | 16 (11.5%) | 8 (12.3%) | 1.000 |
| Vomitting, n(%) | 12 (5.88%) | 8 (5.76%) | 4 (6.15%) | 1.000 |
| Any lower adverse GI effects, n(%) | 136 (66.7%) | 85 (61.2%) | 51 (78.5%) | 0.022 |
| Number lower adverse GI effects | 1.00 [0.00;2.00] | 1.00 [0.00;1.50] | 1.00 [1.00;2.00] | 0.072 |
| Any upper adverse GI effects, n(%) | 186 (91.2%) | 131 (94.2%) | 55 (84.6%) | 0.046 |
| Number upper adverse GI effects | 1.00 [1.00;2.00] | 1.00 [1.00;1.50] | 1.00 [1.00;2.00] | 0.734 |
| Upper and lower adverse GI effects, n(%) | 119 (58.3%) | 77 (55.4%) | 42 (64.6%) | 0.275 |
| Only lower adverse GI effects, n(%) | 68 (33.3%) | 54 (38.8%) | 14 (21.5%) | 0.022 |
| Only upper adverse GI effects, n(%) | 17 (8.33%) | 8 (5.76%) | 9 (13.8%) | 0.094 |
| Number total adverse GI effects | 2.00 [1.00;3.00] | 2.00 [1.00;3.00] | 2.00 [2.00;3.00] | 0.147 |

Table S1. Demographic, anthropomorphic, haematinic and self-reported oral iron gastrointestinal tolerability profile of adult, pre-menopausal women with and without ferritin <12µg/L and a self-reported intolerance to oral iron.

| | **Participants with low iron N=128** | **Excluded from the RCT N=69** | **Included in the RCT N=59** | **P value** |
|---|---|---|---|---|
| Age, years | 36.6 (10.2) | 37.8 (9.35) | 35.2 (11.0) | 0.147 |
| History of iron deficiency, n(%) | 13 (10.2%) | 2 (2.90%) | 11 (18.6%) | 0.008 |
| History of anaemia, n(%) | 17 (13.3%) | 5 (7.25%) | 12 (20.3%) | 0.056 |
| History of iron deficiency or anaemia, n(%) | 18 (14.1%) | 5 (7.25%) | 13 (22.0%) | 0.032 |
| Weight, kg | 71.3 [60.6;79.7] | 70.8 [61.4;78.6] | 73.0 [59.8;82.1] | 0.899 |
| Height, m | 1.65 (0.06) | 1.65 (0.06) | 1.64 (0.06) | 0.558 |
| BMI, kg/m$^2$ | 25.9 [22.2;29.3] | 25.8 [23.0;29.0] | 26.3 [21.9;30.4] | 0.804 |
| Temp, °C | 36.2 (0.46) | 36.2 (0.45) | 36.2 (0.48) | 0.811 |
| SBP, mmHg | 109 [102;117] | 108 [102;116] | 110 [102;118] | 0.303 |
| DBP, mmHg | 73.9 (10.0) | 73.8 (10.6) | 74.0 (9.35) | 0.882 |
| HR, bpm | 70.0 [65.0;77.0] | 70.0 [65.0;78.0] | 71.0 [65.5;76.0] | 0.738 |
| Smoking status: | | | | 0.142 |
| Never smoked, n(%) | 84 (65.6%) | 44 (63.8%) | 40 (67.8%) | |
| Previous smoker, n(%) | 30 (23.4%) | 20 (29.0%) | 10 (16.9%) | |
| Current smoker, n(%) | 14 (10.9%) | 5 (7.25%) | 9 (15.3%) | |
| Alcohol consumption, units/week | 2.48 (2.77) | 2.34 (2.49) | 2.65 (3.08) | 0.542 |
| Depot contraceptive, n(%) | 4 (3.12%) | 1 (1.45%) | 3 (5.08%) | 0.334 |
| Patch or ring contraceptive, n(%) | 6 (4.69%) | 4 (5.80%) | 2 (3.39%) | 0.686 |
| Oral contraceptive, n(%) | 22 (17.2%) | 11 (15.9%) | 11 (18.6%) | 0.866 |
| Serum Iron, µmol/L | 12.7 [7.18;17.1] | 13.3 [7.20;21.4] | 12.7 [7.20;16.6] | 0.845 |
| Total iron binding concentration, µmol/L | 64.0 (8.96) | 62.3 (9.79) | 64.7 (8.58) | 0.293 |
| Transferrin saturation, % | 21.2 [9.85;29.0] | 24.4 [9.00;32.7] | 21.2 [10.0;26.6] | 0.531 |
| Ferritin, µg/L | 11.5 [7.00;15.8] | 13.0 [8.00;18.0] | 9.00 [6.00;14.0] | 0.020 |
| White cell count, $10^{-9}$/L | 5.58 [4.55;6.39] | 5.07 [4.59;6.36] | 5.96 [4.55;6.56] | 0.332 |
| Red cell count, $10^{-12}$/L | 4.41 [4.18;4.66] | 4.46 [4.24;4.76] | 4.38 [4.18;4.60] | 0.404 |
| Haemoglobin, g/dL | 12.8 [11.9;13.6] | 13.0 [12.6;13.7] | 12.1 [11.5;13.4] | 0.001 |
| Haematocrit, L/L | 0.38 [0.36;0.41] | 0.39 [0.37;0.41] | 0.38 [0.36;0.41] | 0.201 |
| Mean cell volume, fL | 88.3 [84.3;90.4] | 89.5 [85.3;92.0] | 87.7 [84.3;89.5] | 0.149 |
| Mean cell Hb, pg | 28.5 [26.6;29.9] | 29.1 [26.6;30.0] | 28.2 [26.7;29.6] | 0.343 |
| Mean cell Hb concentration, g/dL | 32.2 (1.14) | 32.2 (1.11) | 32.1 (1.16) | 0.725 |
| Red cell distribution width, % | 13.6 [13.0;14.7] | 13.3 [12.9;14.4] | 13.7 [13.1;14.7] | 0.233 |
| Platelets, $10^{-9}$/L | 278 [243;323] | 282 [246;310] | 278 [243;338] | 0.862 |
| Neutrophils, $10^{-9}$/L | 3.20 [2.44;4.11] | 2.97 [2.50;4.01] | 3.52 [2.43;4.25] | 0.347 |
| Lymphocytes, $10^{-9}$/L | 1.67 [1.33;1.88] | 1.60 [1.23;1.82] | 1.73 [1.38;1.90] | 0.262 |
| Monocytes, $10^{-9}$/L | 0.44 [0.36;0.52] | 0.44 [0.37;0.52] | 0.44 [0.36;0.52] | 0.955 |
| Eosinophils, $10^{-9}$/L | 0.12 [0.06;0.25] | 0.12 [0.05;0.23] | 0.12 [0.07;0.24] | 0.628 |
| Basophils, $10^{-9}$/L | 0.02 [0.02;0.04] | 0.02 [0.02;0.04] | 0.02 [0.02;0.04] | 0.904 |

| | | | | |
|---|---|---|---|---|
| HxGID | 12 (9.38%) | 6 (8.70%) | 6 (10.2%) | 1.000 |
| Constipation, n(%) | 100 (78.1%) | 56 (81.2%) | 44 (74.6%) | 0.494 |
| Abdominal pain, n(%) | 8 (6.25%) | 5 (7.25%) | 3 (5.08%) | 0.725 |
| Nausea, n(%) | 78 (60.9%) | 35 (50.7%) | 43 (72.9%) | 0.017 |
| Abdominal pain or nausea, n(%) | 54 (42.2%) | 25 (36.2%) | 29 (49.2%) | 0.195 |
| Indigestion, n(%) | 41 (32.0%) | 18 (26.1%) | 23 (39.0%) | 0.171 |
| Heartburn, n(%) | 4 (3.12%) | 3 (4.35%) | 1 (1.69%) | 0.624 |
| Eructation, n(%) | 18 (14.1%) | 7 (10.1%) | 11 (18.6%) | 0.261 |
| Diarrhoea, n(%) | 14 (10.9%) | 7 (10.1%) | 7 (11.9%) | 0.979 |
| Vomitting, n(%) | 6 (4.69%) | 4 (5.80%) | 2 (3.39%) | 0.686 |
| Any lower adverse GI effects, n(%) | 91 (71.1%) | 45 (65.2%) | 46 (78.0%) | 0.164 |
| Number lower adverse GI effects | 1.00 [0.00;2.00] | 1.00 [0.00;1.00] | 1.00 [1.00;2.00] | 0.034 |
| Any upper adverse GI effects, n(%) | 115 (89.8%) | 63 (91.3%) | 52 (88.1%) | 0.766 |
| Number upper adverse GI effects | 1.00 [1.00;2.00] | 1.00 [1.00;2.00] | 1.00 [1.00;1.00] | 0.035 |
| Upper and lower adverse GI effects, n(%) | 79 (61.7%) | 40 (58.0%) | 39 (66.1%) | 0.447 |
| Only lower adverse GI effects, n(%) | 37 (28.9%) | 24 (34.8%) | 13 (22.0%) | 0.164 |
| Only upper adverse GI effects, n(%) | 12 (9.38%) | 5 (7.25%) | 7 (11.9%) | 0.556 |
| Number total adverse GI effects | 2.00 [1.00;3.00] | 2.00 [1.00;3.00] | 2.00 [2.00;3.00] | 0.463 |

Table S2. Comparison of demographic, anthropomorphic, haematinic and self-reported oral iron gastrointestinal tolerability profile of adult, pre-menopausal women with and without ferritin <30μg/L in the screening study (n=128) and the prospective treatment study (n=59).

| | **All Patients N=59** | **IWP 14mg N=18** | **IWP 25mg N=21** | **IWP 50mg N=20** |
|---|---|---|---|---|
| Abdominal pain | 1.25 (0.68) | 1.17 (0.51) | 1.38 (0.86) | 1.20 (0.62) |
| Heartburn | 1.19 (0.92) | 1.00 (0.00) | 1.29 (1.10) | 1.25 (1.12) |
| Reflux | 1.32 (0.95) | 1.28 (0.83) | 1.43 (1.21) | 1.25 (0.79) |
| "Sucking" feeling | 1.32 (0.90) | 1.50 (0.92) | 1.24 (0.62) | 1.25 (1.12) |
| Nausea / vomiting | 1.19 (0.73) | 1.06 (0.24) | 1.10 (0.30) | 1.40 (1.19) |
| Rumbling | 1.32 (0.95) | 1.39 (1.04) | 1.24 (0.62) | 1.35 (1.18) |
| Bloating / gas | 1.73 (1.13) | 2.00 (1.46) | 1.57 (0.98) | 1.65 (0.93) |
| Burping / belching | 1.22 (0.70) | 1.11 (0.47) | 1.38 (0.86) | 1.15 (0.67) |
| Flatulence | 1.44 (1.05) | 1.94 (1.55) | 1.29 (0.78) | 1.15 (0.49) |
| Constipation | 1.42 (0.86) | 1.56 (0.92) | 1.43 (0.93) | 1.30 (0.73) |
| Diarrhoea | 1.20 (0.69) | 1.22 (0.65) | 1.24 (0.77) | 1.15 (0.67) |
| Loose stools | 1.19 (0.66) | 1.22 (0.65) | 1.19 (0.68) | 1.15 (0.67) |
| Hard stools | 1.22 (0.65) | 1.50 (0.99) | 1.19 (0.51) | 1.00 (0.00) |
| Defaecation urgency | 1.08 (0.38) | 1.17 (0.51) | 1.10 (0.44) | 1.00 (0.00) |
| Incomplete emptying | 1.32 (0.84) | 1.56 (1.10) | 1.33 (0.86) | 1.10 (0.45) |
| Overall GSRS gut symptom score | 19.4 (7.05) | 20.7 (7.39) | 19.4 (6.67) | 18.4 (7.31) |

Table S3. Baseline Gastrointestinal Symptom Rating Scale (GSRS) gut symptoms score following washout of 9.8 days of participants with ferritin <30µg/L and self-reported gastrointestinal intolerance to oral iron, randomised to three different daily elemental iron doses of IWP (14mg, 25mg, 50mg).

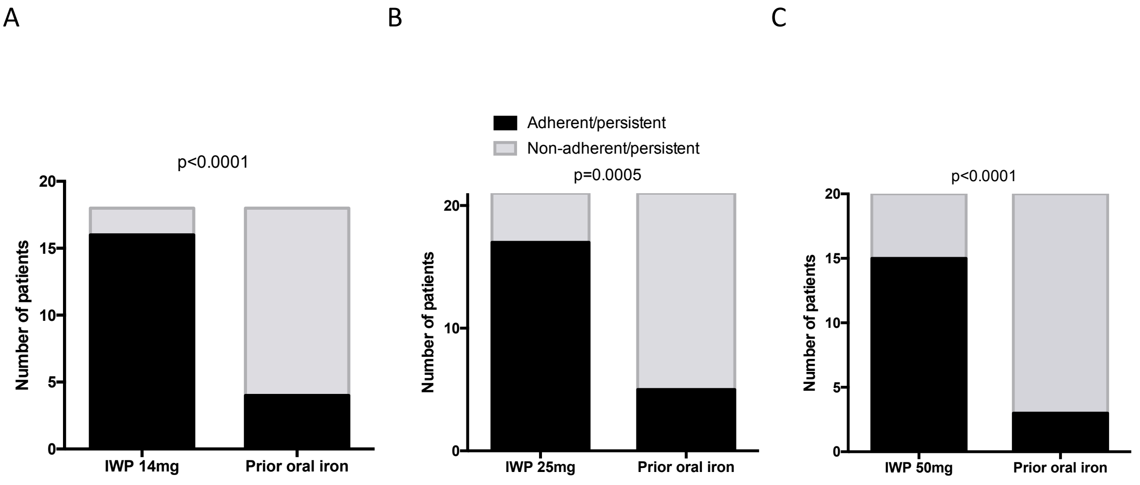


Figure S1. Overall adherence/persistence with IWP amongst 59 women with a history of intolerance to oral iron and with low iron, moderate to severe iron deficiency or iron deficiency anaemia. Data show good adherence/persistence, compared to the previous oral iron according to the random allocation to daily dose group 14mg (A), 25mg (B) and 50mg.

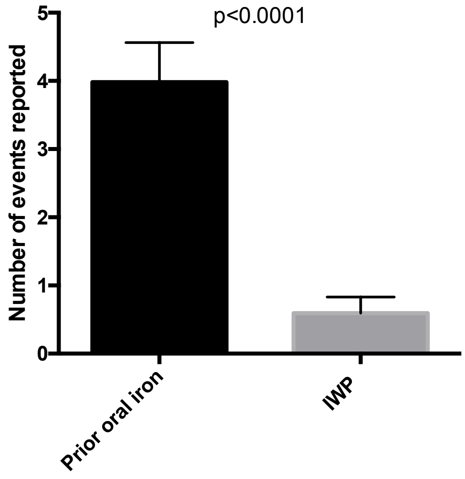


Figure S2. Number of elicited adverse GI events reported with previous oral iron product with IWP amongst 59 women with a history of intolerance to oral iron with low iron, moderate to severe iron deficiency or iron deficiency anaemia.

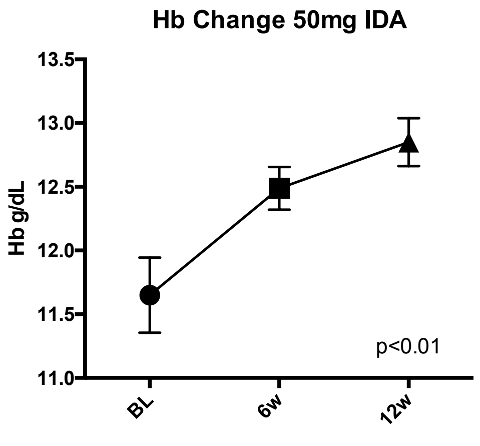


Figure S3. Overall adherence/persistence with IWP amongst 59 women with a history of intolerance to oral iron with low iron, moderate to severe iron deficiency or iron deficiency anaemia, compared to the previous oral iron according to the random allocation to daily dose group 14mg (A), 25mg (B) and 50mg.

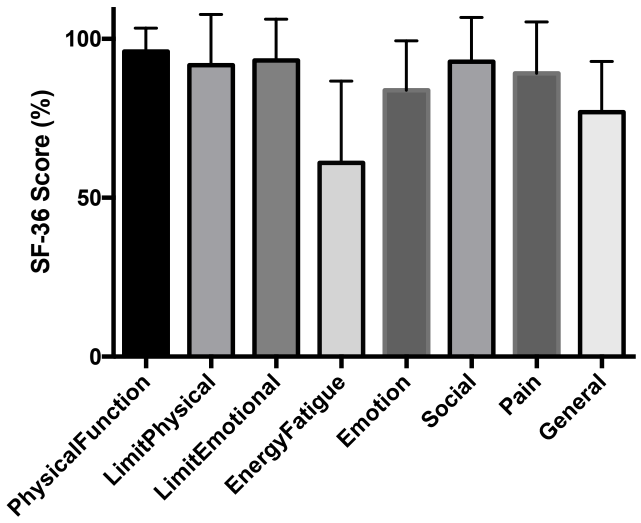


Figure S4. Detailed SF-36 data broken down according to the 8-health related quality of life concepts: physical functioning, bodily pain, role limitations due to physical health problems, role limitations due to personal or emotional problems, emotional well-being, social functioning, energy/fatigue, and general health perceptions. Using ANOVA, there were significant differences across the domains ($P<0.0001$) and SF-36 Energy/Fatigue domain scores in this population ($60 \pm 4\%$) were significantly impaired compared to all the other domain scores at baseline (all $P<0.001$).
